# Genetic Risk of Axonal Neuropathy Following Infection

**DOI:** 10.1101/2024.10.04.24314535

**Authors:** J. Robert Harkness, John H. McDermott, Shea Marsden, Peter Jamieson, Kay A. Metcalfe, Naz Khan, William L. Macken, Robert D.S. Pitceathly, Christopher J. Record, Reza Maroofian, Kloepa Kloepas, Ataf Sabir, Lily Islam, Saikat Santra, Enise Avci Durmusalioglu, Tahir Atik, Esra Isik, Ozgur Cogulu, Jill E. Urquhart, Glenda M. Beaman, Leigh A. Demain, Adam Jackson, Alexander J.M. Blakes, Helen M. Byers, Hayley Bennett, Wei-Hsiang Lin, Antony Adamson, Sanjai Patel, Wyatt W. Yue, Robert W. Taylor, Janine Reunert, Thorsten Marquardt, Rebecca Buchert-Lo, Tobias Haack, Heike Losch, Lukas Ryba, Petra Lassuthova, Radka Valkovičová, Jana Haberlová, Barbora Lauerová, Eva Trúsiková, Kiran Polavarapu, Ozge Aksel Kilicarslan, Hanns Lochmüller, Mina Zamani, Niloofar Chamanrou, Gholamreza Shariati, Saeid Sadeghian, Reza Azizimalamiri, Sateesh Maddirevula, Muhammad AlMuhaizea, Fowzan S. Alkuraya, Rita Horvath, Serdal Gungor, Emma Wakeling, Adnan Manzur, Pinki Munot, Rachael Matthews, Siddharth Banka, Mary M. Reilly, Daimark Bennett, Raymond T. O’Keefe, William G. Newman

## Abstract

**Background:** Why some individuals experience severe neuropathy following infection is unknown. Nucleocytoplasmic trafficking (NCT) is an essential process in nucleated cells, and its disruption has been implicated in many neurodegenerative conditions including amyotrophic lateral sclerosis (ALS) and frontotemporal dementia.

**Methods:** We performed genomic and clinical studies in 24 individuals from 12 families with acute onset axonal neuropathy. Genetic variants were characterized by thermal stability and enzymatic assays using recombinantly expressed protein. Protein localization was determined in patient fibroblasts using immunofluorescence following heat or oxidative stress. A humanized *Drosophila* model was generated to determine the effect of stress on *in vivo* function.

**Results:** We identified deleterious biallelic variants in human *RCC1*, encoding a GTP exchange factor essential in maintaining Ran GTPase-dependent NCT function. Clinical presentations ranged from a rapidly progressive, fatal axonal neuropathy with encephalopathy to a mild motor neuropathy resulting in impaired walking. In most patients (n=22/24), neurological presentation was secondary to infection, resulting in prior diagnosis of Guillain-Barré syndrome (GBS) in 13. The efficiency of cellular Ran GDP-GTP exchange and the thermal stability of Rcc1 protein was reduced by disease-associated variants. Heat shock or oxidative stress revealed defects in Ran nuclear localization, impaired NCT, and TDP-43 mislocalization in patient fibroblasts. Disease associated variants were unable to rescue the thermosensitive phenotype of a *rcc1* deficient hamster cell line. *RCC1 Drosophila* models revealed a fatal intolerance to oxidative stress.

**Conclusion:** We describe a novel autosomal recessive acute onset axonal neuropathy triggered by infection caused by biallelic *RCC1* variants, which mimics GBS and has important mechanistic overlap with ALS.

## Introduction

Eukaryotic cells can be distinguished from their prokaryotic ancestors by the presence of a nuclear membrane which separates genetic material from the rest of the cell. A tightly regulated process manages the trafficking of molecules across this membrane via nuclear pore complexes (NPCs).^1^ Export of nuclear cargoes via the NPC requires the formation of exportin-cargo complexes, which are assembled in a Ran-guanosine triphosphate (Ran-GTP) dependent process (**Figure 1**).^2^ A homeostatic Ran-GTP gradient is maintained by the chromatin bound Rcc1 protein which acts as a guanine exchange factor (GEF) for Ran, catalyzing guanosine diphosphate (GDP) to GTP exchange.^3,4^ The proteins involved in this nuclear trafficking pathway are highly conserved throughout eukaryotic evolution, and deleterious variants in several NPC genes are implicated in Mendelian disease.^1,5,6^

**Figure 1.**
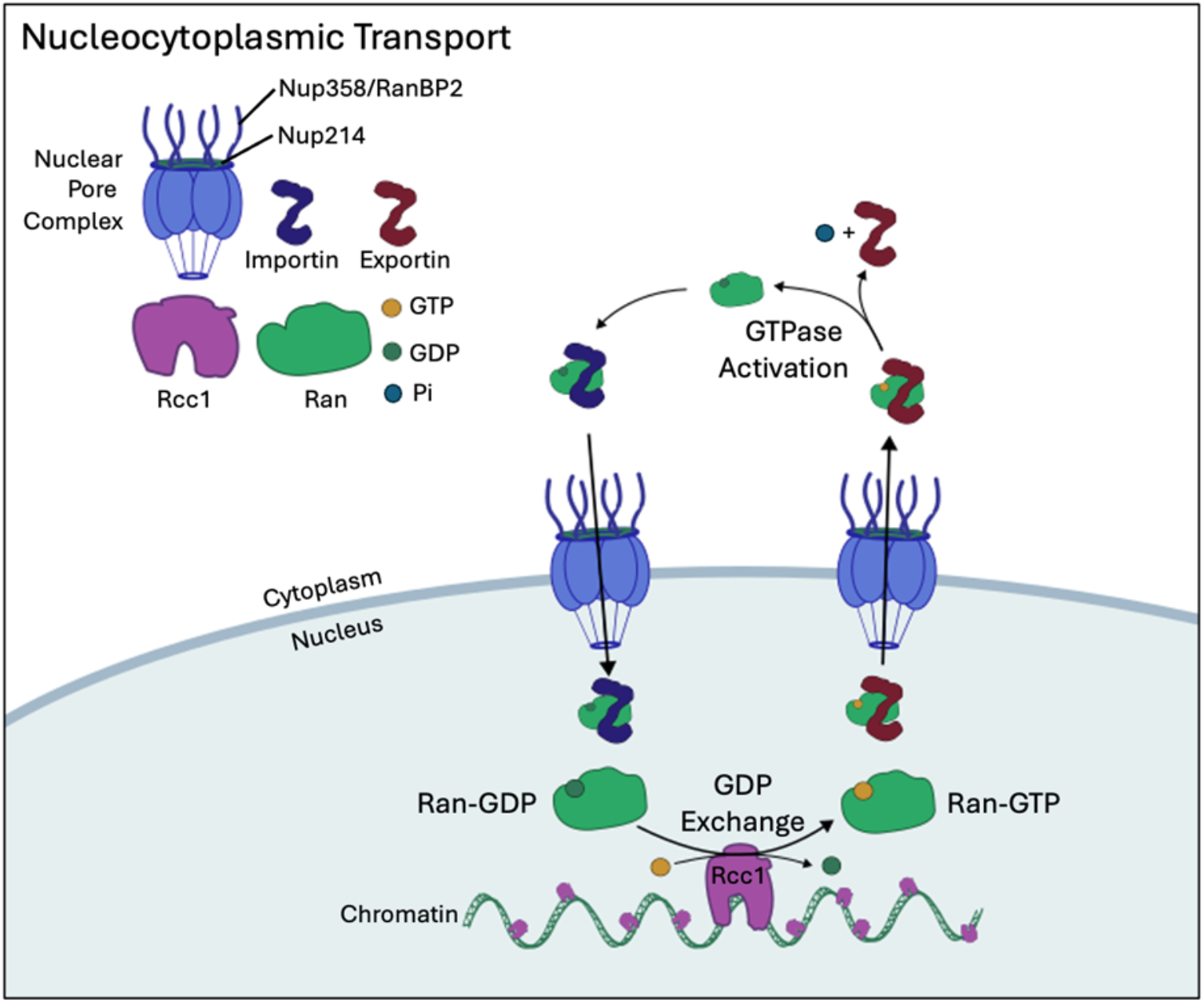
Nucleocytoplasmic transport. Transport of cargoes between the cytoplasm and the nucleus is facilitated by Ran, a small nuclear GTPase protein. In the nucleus, Ran is loaded with GTP by Rcc1, the only known GTP exchange factor (GEF). Rcc1 binds to chromatin, anchoring Ran GDP-GTP exchange in the nucleus. Ran-GTP then associates with an exportin complexed with cargo to be exported from the nucleus. The Ran-GTP-exportin complex passes though the nuclear pore, where Ran-GTPase Activating Proteins (Ran-GAPs) such as RanBP2 catalyze GTP hydrolysis by Ran, facilitating Ran-Exportin-Cargo complex dissociation. Ran, now GDP-bound is transported back into the cytoplasm in complex with an importin, and nuclear-bound cargo, for further cycles of Rcc1-dependent GDP-GTP exchange. Pi = inorganic phosphate. GDP = Guanosine diphosphate. GTP = guanosine triphosphate.

Nucleocytoplasmic trafficking (NCT) is an essential process in nucleated cells, facilitating transport of inbound genomic and cell cycle regulatory factors and export of mRNA transcripts. Identification of patient cohorts with inherited defects in NCT therefore provides an opportunity to investigate the role of this process in health and disease. Characterizing the clinical and cellular profiles of these cohorts could also aid understanding of the etiologic links between impaired NCT and common disorders such as amyotrophic lateral sclerosis (ALS) and frontotemporal dementia (FTD).^7–9^ These insights may also contribute towards the development of therapeutic strategies which rescue or augment NCT function.^10,11^

Deleterious variants in two genes, *RANBP2* and *NUP214*, which code for proteins in the outer ring of the NPC demonstrate how abnormally attenuated NCT in response to infection can lead to human disease.^12,13^ Individuals with damaging variants in *RANPB2* and *NUP214* are predisposed to infection-induced acute-onset encephalopathy. Why some patients are susceptible to severe neurological complications from infection is not well understood, though familial cases provide a unique opportunity to investigate this susceptibility.

Here, we report the discovery of 24 individuals from 12 families with deleterious biallelic variants in *RCC1*. Affected individuals presented with an acute-onset neuropathy following infection which was distinguished by variable weakness, hypotonia and areflexia. We highlight NCT as a vital mechanism that is susceptible to disruption following cellular stress resulting in defects in protein homeostasis.

## Methods

### Patients

Through international collaboration, we identified 24 individuals from 12 families in whom rare and predicted protein damaging, biallelic *RCC1* (NM_001381865.2) variants were identified. Following identification of the index family (Family 1), further participants were ascertained through GeneMatcher, the 100,000 Genomes Project and SOLVE-RD.^14,15^ Details of the clinical findings are provided in **Table S1** in the supplementary appendix. Written informed consent was obtained from all persons in the study (or from their parents or guardians) in accordance with the Declaration of Helsinki protocols, and our experimental protocols were approved by local institutional review boards (IRAS 64321). The authors vouch for the accuracy and completeness of the data presented in this report.

### Genetic Analyses

Exome or genome sequencing was performed with germline DNA from at least one affected individual in each family. Sequence data were filtered based on variant frequency and computer modelling tools were used to predict variant pathogenicity. Variants were tested for segregation in affected and unaffected family members via Sanger sequencing. Full details are provided in the methods section of the supplementary appendix.

### Protein Modelling

Three-dimensional structures of wild-type (WT) Rcc1 protein in the context of the Ran-Rcc1 complex (PDB ID: li2m) and the Rcc1-nucleosome complex (PDB ID: 3mvd) were used to map each variant site.

### Rcc1 Activity and Stability

Recombinant 6xHis-tagged wildtype and mutant Rcc1 proteins were expressed from pET-28a vectors in Rosetta DE3 E. coli cultures. His-tagged proteins were purified by gravity flow using Nickel His-Select Agarose beads (Sigma, UK). The activities of recombinant wild type and mutant Rcc1 proteins were assessed via a guanine exchange factor (GEF) assay, establishing the ability of recombinant Rcc1 to facilitate guanine exchange with Ran GTPase. The thermal stability of the recombinant Rcc1 proteins, with and without recombinant Ran, was examined via a thermal shift assay. Fluorescence was measured using a StepOnePlus™ Real-Time PCR System (Applied Biosystems) after each 1 % increase in temperature between 25 °C and 99 °C. A detailed description of the GEF and thermal shift assays are provided in the supplementary appendix.

### Cell Studies

Primary skin fibroblast cell lines from anonymized healthy controls and patients were cultured. Rcc1 and TDP-43 protein expression was quantified via western blotting. For functional studies, cellular stress was induced by means of heat shock (43°C) for 4-20 hours, or with the use of hydrogen peroxide (H_2_O_2_) at a concentration of 200μM or 500μM. Localization of Rcc1, Ran and TDP-43 proteins was investigated via immunofluorescence, and the Ran nuclear:cytoplasmic ratio was quantified under control and stressed conditions. Rescue studies were undertaken by transfecting the tsBN2 (Rcc1^Ser256Phe^/^Ser256Phe^) cell line with constructs containing mScarlet-I-tagged wild type or mutant *RCC1*. Detailed descriptions of the methods used for cell culture, immunofluorescence, and rescue studies are provided in the supplementary appendix.

### *Drosophila* Studies

Fly strains were reared on standard yeast-sugar-agar media at 25 °C. For functional studies, cellular stress was induced with paraquat (PQ) at 20-50 μM (Sigma). Detailed descriptions of the methods used for generating CRISPR-Cas9 knock-ins, immunofluorescent imaging and functional analysis, together with details of *Drosophila* genotypes, can be found in the supplementary appendix.

## Results

### Genetic association between rare *RCC1* variants and axonal neuropathy

Patients with damaging biallelic *RCC1* missense variants from 12 families (**Supplementary Data Table S1**) presented with acute onset weakness and hypotonia between the ages of 7 months and 11 years of age (median 18 months). Almost all individuals (22/24) developed symptoms in the days following a mild and often undefined infection, resulting in a differential diagnosis of Guillain-Barré syndrome (GBS) in many (13/24) of the patients. A variety of precipitating illnesses were reported, including respiratory tract infections (9/24), ear infections (1/24), diarrhea (2/24), and non-specific fever (5/24). One patient presented with symptoms following a raised temperature in the days after their 12-month immunizations. As well as weakness and hypotonia, most patients presented with absent reflexes (20/24) and bulbar involvement (17/24). Most patients required intensive care unit support on admission, with respiratory muscle involvement and ventilatory support. A detailed case report for each patient is available upon request.

Nerve conduction studies in affected patients identified patterns consistent with axonal motor neuropathy, with or without sensory nerve involvement. Brain MRI identified cerebral atrophy in eight of the fourteen individuals who underwent assessment (**Figure 2**). Brain imaging was often normal immediately following presentation, but abnormalities became apparent with serial imaging over the course of several months with a progressive loss of brain parenchyma, often with a fronto-temporal predominance. There were no consistent electroencephalogram findings in affected individuals, whilst six of the patients had raised protein in cerebrospinal fluid in the acute phase of presentation. Several of the patients were treated with either intravenous immunoglobulin (IVIg) or plasma exchange, but these therapies did not result in symptomatic improvement. Genetic testing identified biallelic *RCC1* missense variants in all families, either in homozygous or compound heterozygous state (**Supplementary Data Table S6**).

**Figure 2.**
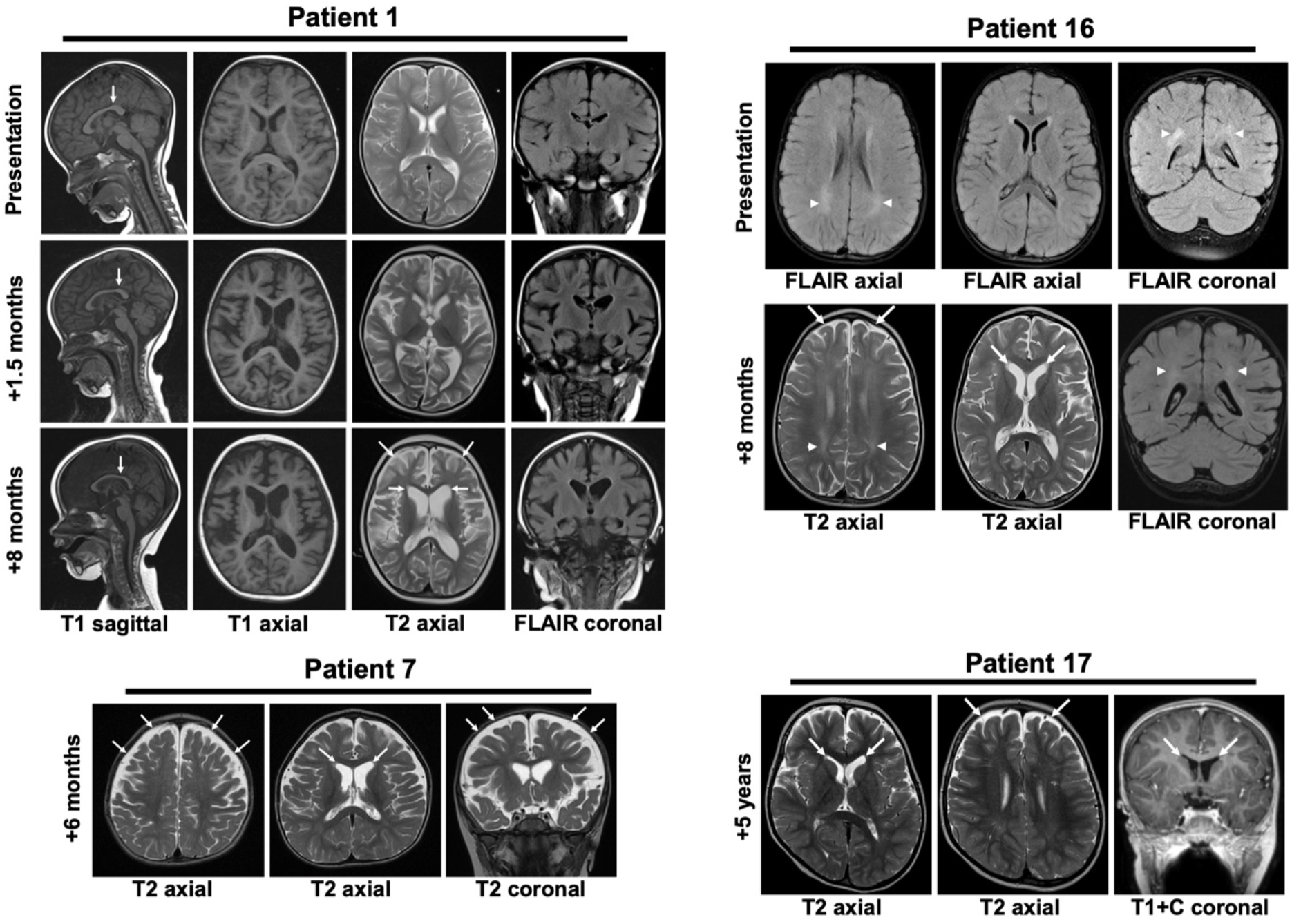
(Selected MRI brain images for PATIENT 1, PATIENT 7, PATIENT 16 and PATIENT 17. Serial scans are shown for PATIENTS 1 and 16. Imaging shows evidence of progressive cortical and white matter atrophy with frontotemporal predominance, causing prominence of the ventricles and extra-axial CSF spaces (arrows) and on some scans there is white matter hyperintensity on T2-weighted imaging (arrowheads). The date the scan was performed is shown in months, with the PATIENT’S presentation as the baseline.

Symptomatic recovery was extremely limited across the cohort, with only one patient (Patient 19) demonstrating complete recovery over the ensuing months. Over half (13/24) of the patients had some limited recovery in their symptoms, but many required long-term ventilatory support with persistent weakness. Recurrent episodes were observed in the majority of patients (15/24) and were also precipitated by periods of illness, with or without fever. These subsequent episodes often led to a severe deterioration in symptoms, and 15 individuals died as a result of their condition between the ages of 11 months and 23 years of age.

### Patient-associated variants alter Rcc1 protein thermal stability and function *in vitro*

The sites of the nine missense variants are distributed across the different domains of the Rcc1 polypeptide, including the protein core, and the known interface with Ran and the nucleosome (**Supplementary Data Figure S2**). To assess the impact of variants on the stability of Rcc1 proteins, we expressed and purified recombinant wild-type and variant versions of the human Rcc1 protein (**Figure 3A, Supplementary Data Figure S3**). A thermal stability assay (TSA) was used to characterize the thermal denaturation profiles of recombinant Rcc1 variants, as the tsBN2 cell line with a Rcc1 p.(Ser256Phe) homozygous variant (**Figure 3A**) displays a thermosensitive phenotype and onset of neurological deficits in patients is associated with fever.^16^ All disease-associated variants displayed reduced thermal stability (**Figure 3B, Supplementary Data Figure S4**). Variability in the stability of Rcc1 was also supported by reduced Rcc1 levels in whole cell lysates from patient fibroblasts (**Supplementary Data Figure S5)**.

**Figure 3.**
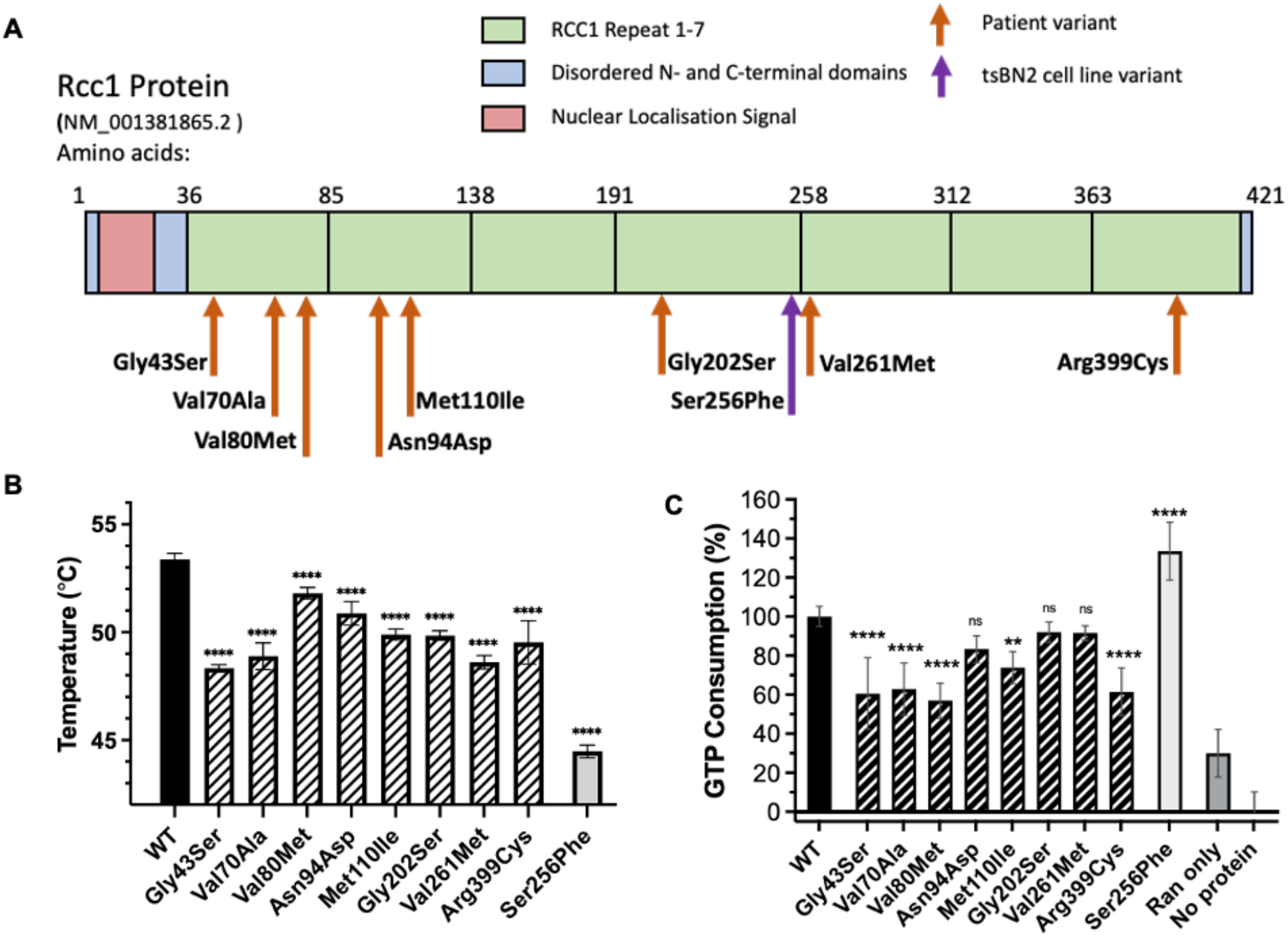
(A) Plot of the Rcc1 protein MANE transcript (ENST00000683442.1; NM_001381865.2) with location of missense variants identified in patients which were expressed recombinantly. Patient variants (orange arrows) are located in five different Rcc1-repeat domains. The Ser256Phe variant (purple arrow) was previously identified in the tsBN2 cell line and confers a temperature sensitive phenotype. (B) Melting temperature (Tm) of recombinant Rcc1 mutant proteins determined via protein thermal stability assay. Data represent mean Tm ± s.d. (n=3). (C) GTP consumption by recombinant Ran GTPase in GTP Exchange Factor (GEF) assay with Rcc1 protein mutants. Data represent mean relative GTP consumption ± s.d. (n=3). Data were subjected to Dunnett’s multiple comparisons test: ns non-significant; ** p<0.01; **** p<0.0001. WT wild-type.

To determine the functional consequence of missense variants on Rcc1 ability to facilitate GDP-GTP exchange with Ran, GEF assays were conducted using recombinantly expressed Rcc1 and Ran proteins. Relative to activity of wild-type (WT) Rcc1, five patient variants (Gly43Ser, Val70Ala, Val80Met, Met110Ile, and Arg399Cys) demonstrated a significant reduction in GTP consumption (56.97 % to 73.82 % of WT activity) (**Figure 3C**). Three variants identified in patients (Asn94Asp, Gly202Ser, and Val261Met) displayed small, statistically insignificant decreases in activity (83.33 % to 93.35 % of WT activity).

### Cell models of *RCC1* variants indicate stress-induced disruption of Ran gradients

To characterize the functional deficits of *RCC1* mutants with greater biological context, we examined Ran localization in primary dermal fibroblasts obtained from six patients. Fibroblasts were cultured under standard conditions (37.0 °C, 5.0 % CO_2_), and stressed by addition of 200-500 µM H_2_O_2_, or by exposing cells to heat shock by incubating at 43 °C for 4-20 hours.

Under stressed conditions, nuclear Rcc1 protein staining decreased and became peri-nuclear in localization (**Figure 4A**). Under standard culture conditions, Ran predominantly localized to the nucleus, where GDP-GTP exchange is performed by Rcc1. However following stress induction, Ran nuclear localization decreased, with accumulation evident around the nuclear envelope in some cells (**Figure 4A, white arrow**). This change in localization indicates defects in the maintenance of the steep Ran-GTP gradients which are essential for maintaining effective transport of cargoes between the nucleus and cytoplasm. To compare between control and patient cell lines, we quantified the ratio of nuclear:cytoplasmic Ran fluorescence intensity (**Figure 4B**). This quantification demonstrated that there was a significantly greater proportion of cellular Ran localization in the nucleus of control cells compared to patient fibroblasts under standard culture conditions. Under conditions of stress, an overall reduction in Ran nuclear staining was observed in both control and patient fibroblast cell lines, though the reduction in patient cells was significantly greater (**Figure 4B**).

**Figure 4.**
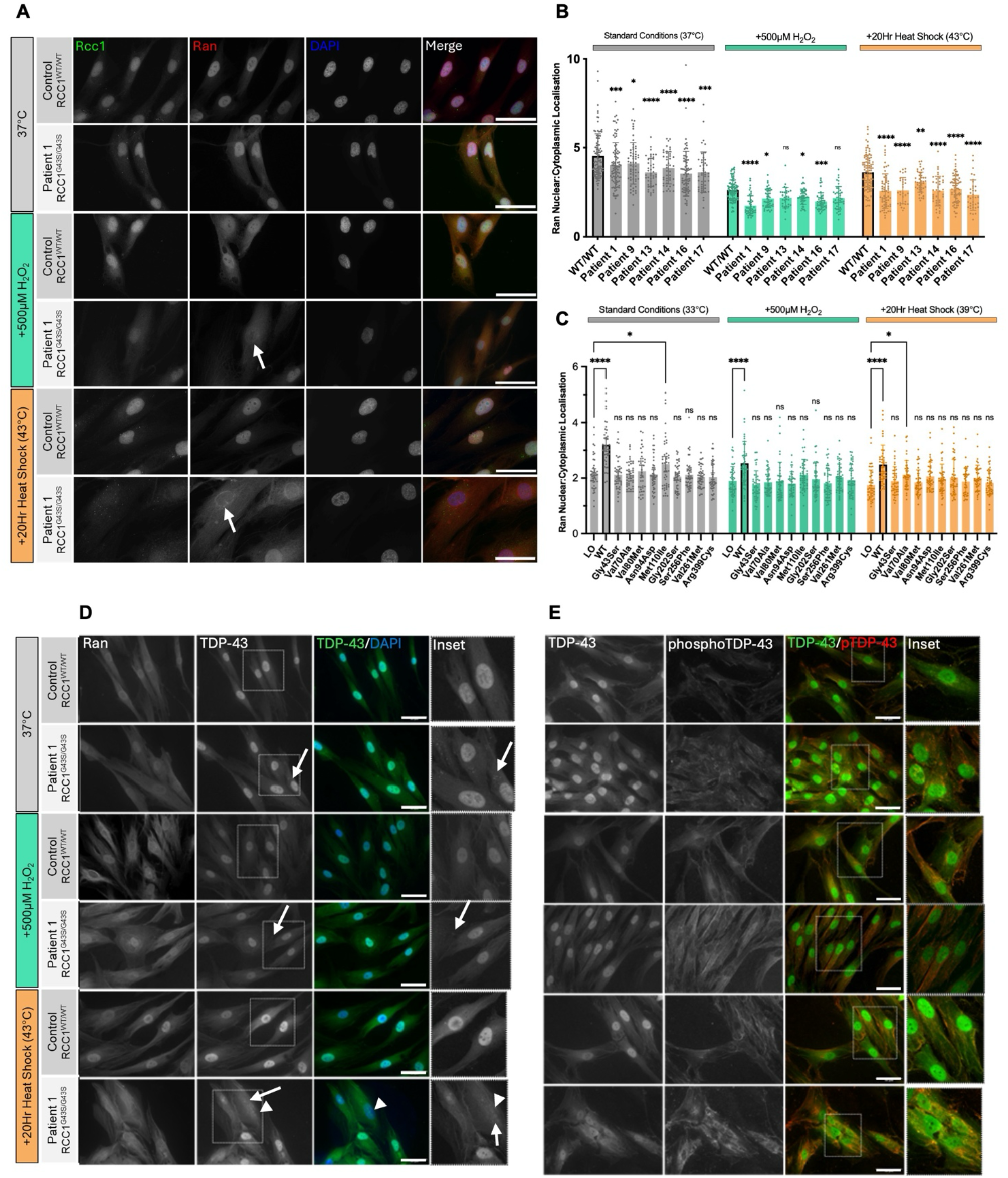
Immunofluorescence analysis of *RCC1* variants in cell models. (A) Immunofluorescence staining of primary human fibroblasts derived from Patient 1 (p.Gly43Ser) under standard culture conditions, and after induction of oxidative or heat stress. Oxidative stress was initiated by addition of 500µM H_2_O_2_ for one hour before fixation. Heat stress was initiated by incubation at 43°C (5.0% CO_2_) for 20 hours before fixation. Cells were stained with antibodies against Rcc1 and Ran, and DAPI. Cells were imaged at 40x using a snapshot microscope. Scale bar 100µm (B) Quantification of Ran nuclear:cytoplasmic ratio in primary patient fibroblasts. Genotypes assessed were Patient 1 (Gly43Ser/Gly43Ser), Patient 9 (Val80Met/Arg399Cys), Patient 13 (Asn94Asp/Asn94Asp), Patient 14 (Met110Ile/Met110Ile), Patient 16 (Arg399Cys/Arg399Cys), and Patient 17 (Met110Ile/Gly202Ser). Bars represent mean ratio of nuclear to cytoplasmic Ran fluorescence value (± 95% confidence interval (CI), n=3), with each point representing measurement from one cell. (C) Quantification of Ran nuclear:cytoplasmic ratio in hamster kidney cell line tsBN2 transfected with human *RCC1* WT and patient variants at standard tsBN2 culture conditions (33°C, 5.0% CO_2_), plus induction of oxidative stress (one hour 500µM H_2_O_2_) and heat stress (39°C). Data represented are mean (± 95% CI, n=3) with each point representing measurement from one cell. Data were subjected to Dunnett’s multiple comparisons test, *p<0.05, **p<0.01, ***p<0.001, ****p<0.0001, ns non-significant. LO lipofectamine only, WT wild-type. (D,E) Immunofluorescence imaging of TDP-43 in fibroblasts from Patient 1. Cells were stained with DAPI, anti-TDP-43, and either anti-Ran (D) or anti-phospho-TDP (p.Ser409/410) (E). Cells were imaged at 40x and 63x (inset). Scale bar 50µm. (D) Culture of patient cells at 43 °C results in TDP-43 localisation to the cytoplasm (arrowhead) in cells where Ran gradients are disrupted. (E) Aggregates of TDP-43 (white arrow) which colocalize with TDP phosphorylated at Ser409/410 are present in patient fibroblasts at 37 °C and 43 °C, with greater TDP phosphorylation in patient cells compared to control under both culture conditions.

To determine if disease associated variants could rescue Rcc1 function, we devised a model examining Ran localization in the tsBN2 hamster kidney cell line.^16^ tsBN2 cells were transfected with wild-type or variant m-Scarlet-tagged *RCC1*, then visualized by immunofluorescence microscopy. Under standard tsBN2 culture conditions (33.0 °C, 5.0 % CO_2_), Ran nuclear localization increased following transfection with WT *RCC1* (**Figure 4C**). All variants except Met110Ile in unstressed conditions and Val70Ala during heat stress displayed no increase in Ran nuclear localization compared to untransfected tsBN2 cells. This failure to restore Ran nuclear localization suggests sustained impairment of GDP-GTP exchange and NCT capacity due to patient associated *RCC1* variants (**Figure 4C**).

Next, we hypothesized that disruption to NCT may cause imbalances in protein processing. Given the predominantly motor neuropathy observed in patients, we stained fibroblasts for TDP-43, a nuclear protein responsible for mRNA chaperoning which is known to aggregate in a range of neurodegenerative diseases.^17^ Under standard culture conditions, TDP-43 localizes predominantly to the nucleus in control fibroblasts. In fibroblasts from Patient 1 TDP-43 also predominantly localizes to the nucleus, however TDP-43 aggregates were identified in the cytoplasm (**Figure 4D**). Following 20-hour heat shock, nuclear staining of TDP-43 decreased compared with control cells. Phosphorylation of TDP-43 at Ser409/410 is a precursor to caspase-dependent cleavage of C-terminal fragments (CTF) which are prone to aggregation in brains of individuals affected by ALS.^18^

Greater levels of phospho-TDP staining were identified in patient fibroblasts following 20 hour heat shock compared to controls (**Figure 4E**) suggesting greater TDP-43 cleavage occurred in patient cells which have defective NCT when exposed to conditions of stress.

### Loss-of-function of *RCC1* sensitizes a *Drosophila* model to environmental stress

To investigate the role of *RCC1* in modulating neuronal function and viability in response to environmental stressors, we perturbed *RCC1* function in an animal model. We engineered a humanized *Drosophila* strain using CRISPR-Cas9, substituting the fly *rcc1* coding sequence, which is essential for viability, with mScarlet-tagged human *RCC1* wild type (hRCC1^wt^) or Gly43Ser (hRCC1^Gly43Ser^).^19^ This engineering placed fluorescently labelled versions of the human gene under transcriptional control of *Drosophila* regulatory elements (**Figure 5A**). We found broad distribution of mScarlet signal throughout the adult brains of flies carrying *hRCC1*^*wt*^ and *hRCC1*^*Gly43Ser*^ (**Figure 5B**), consistent with previous reports of roles for *RCC1* in neuronal development and function.^19,20^ To characterize the phenotypic effects of the Rcc1 Gly43Ser variant, we analysed the viability of biallelic *hRCC1 Drosophila* knock-in strains with or without environmental stress. Both *hRCC1*^*wt*^ and *hRCC1*^*Gly43Ser*^ were capable of developing to adulthood in the absence of stress, in keeping with previous observations that fly *rcc1* shares equivalent functions to mammalian *Rcc1*.^21^ We stressed animals by feeding with paraquat (PQ), a redox-cycling agent that induces oxidative stress and neurotoxicity.^22,23^ PQ sensitivity is a robust indicator of sensitivity to oxidative stress in *Drosophila*.^24,25^ *hRCC1*^*Gly43Ser*^ homozygotes were intolerant to PQ compared to *hRCC1*^*wt*^ flies (**Figure 5C**), indicating that *hRCC1*^*Gly43Ser*^ has diminished resistance to oxidative stress.

**Figure 5.**
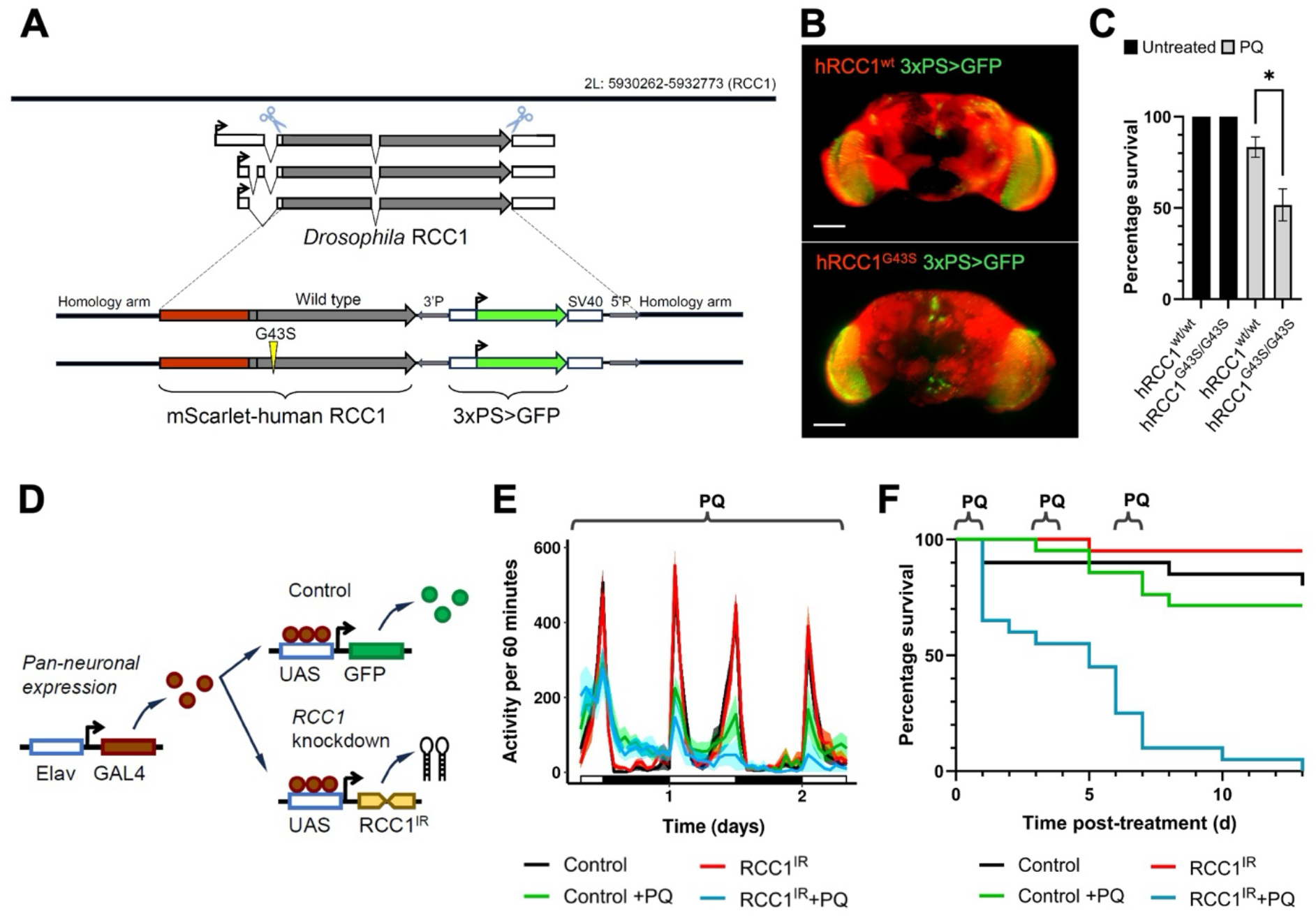
Modelling *RCC1* loss of function in *D. melanogaster*. **A**, Genomic region showing exon-intron structure for *Drosophila* RCC1. Nucleotide coordinates on 2L chromosome are as indicated. Grey shading represents coding regions, unfilled boxes represent untranslated regions with arrows indicating direction of transcription. Cut marks indicate points of insertion of engineered constructs (below), flanked by homology arms, containing mScarlet-RCC1 coding sequence, and 3xPS>GFP marker. **B**, Maximal projection Lightsheet microscope images through a medial region of the adult brain from hRCC1^wt^ and hRCC1^G43S^ flies showing broad expression of mScarlet-labelled hRCC1 expression (red). 3xPS>GFP marker (green) labels photoreceptor neurons and ocelli. Scale bar 100μm. **C**, Percentage survival of animals fed 30 mM paraquat (PQ) prior to eclosion. Survival of PQ-treated *hRCC1*^*Gly43erS*^ animals was significantly reduced compared to hRCC1^wt^ control. Graph shows mean percentage survival ± SEM, n=6 (*, Student’s t-test P=0.012). **D**, Schematic outlining GAL4-UAS expression of control (*UAS-GFP*) or *UAS-RCC*^*IR*^ pan-neuronally, using *elav-GAL4*. **E**, Control and *RCC*^*IR*^ flies show a typical bimodal pattern of locomotor activity during the light-dark cycle (see unfilled and filled x-axis rectangles, respectively). However, *RCC*^*IR*^ flies (blue line) showed increasingly diminishing activity upon chronic exposure to 30 mM PQ. Mean activity ±SEM (n=3) is shown with line and shading; genotypes are as indicated in color key. **F**, Survival curve (Kaplan-Meier) of adult flies exposed to alternating periods of 20 mM PQ treatment and recovery at indicated timepoints. *RCC*^*IR*^ animals show greatly reduced survival (median survival 5 days), compared to treated control or untreated animals. P<0.0001 Log-rank Mantel-Cox test.

*Drosophila rcc1* loss-of-function mutations result in neuronal defects. Therefore, we tested for synthetic lethal interactions between oxidative stress and *RCC1* loss-of-function specifically in the nervous system. We expressed *RCC1* inverted repeat constructs (*RCC1*^*IR*^) using the GAL4-UAS system to induce pan-neuronal RNA interference (**Figure 5D**). *RCC1*^*IR*^ flies exposed to chronic low dose PQ treatments exhibited increasingly diminishing locomotive activity consistent with motor neuropathy in patients (**Figure 5E**). To mimic the escalating response to environmental stress seen in patients, we treated animals with alternating periods of acute PQ-induced stress followed by PQ withdrawal and recovery. Similar to the *hRCC1* knock-ins, we found that flies with neuronal *RCC1*^*IR*^ showed decreased survival in response to PQ-induced stress compared to controls (**Figure 5F**). With more intensive PQ treatments, median survival was 3.6 days for neuronal *RCC1*^*IR*^, compared to 7.3 days for control flies (n=4, P<0.0001 Log-rank Mantel-Cox test). Together, these data indicate that loss-of-function of *RCC1* in the nervous system sensitizes animals to environmental stress leading to locomotion defects and early demise.

## Discussion

Our work demonstrates the clinical significance of disrupted NCT in humans. We identified 24 individuals from 12 families with deleterious, recessive variants in *RCC1*, a gene encoding a protein essential in maintaining a homeostatic Ran-GTP gradient, critical for normal NCT.

Defective nuclear trafficking, precipitated by cellular stress, underlies the rapid and profound neurological impairment in our patients. The missense variants identified reduced the thermal stability of the Rcc1 protein, with some variants associated with decreased GTPase activity *in vitro*. Patient fibroblast studies demonstrated that under standard culture conditions the variants do not impact Ran or Rcc1 localization. However, when patient fibroblasts were cultured at non-permissive temperatures or when oxidative stress was induced by exposure to hydrogen peroxide, Ran-GTP gradients were disrupted with cytoplasmic accumulation and decreased levels in the nuclear compartment. Fly studies indicated that, when stressed, altered Rcc1 results in diminished locomotor function and early death. Moreover, we observed mislocalization of TDP-43 in patient fibroblasts, with greater phosphorylation of TDP-43 compared to controls, a phenomenon observed in the tissues of individuals with both familial and sporadic ALS and frontotemporal dementia (FTD).^17^

Our clinical characterization of the patients included in this cohort demonstrates a consistent phenotype of a severe and rapid onset motor neuropathy, precipitated by periods of fever or infection. In the absence of a diagnosis, the temporal association between infection and symptom onset led to the diagnosis of GBS in many of the patients. However, whilst the natural history of GBS tends towards recovery, almost all of the patients described in this cohort had a limited recovery, indicating permanent neuronal damage.^26^ In addition, unlike GBS, there was central nervous system involvement in eight patients in this cohort with progressive cerebral atrophy with frontotemporal predominance observed in serial brain imaging.

Although most patients were severely affected, we observed some phenotypic heterogeneity, most notably in Family 9. Patient 20 was the least severely affected member of this cohort, developing a single episode of lower-limb weakness with full recovery whilst his sibling, Patient 19, died after a single episode following an infection. If such variability was observed between families this may be explained, in part, by the observed differential impact of *RCC1* variants on protein thermostability. However, this mechanism is insufficient to explain phenotypic variation within a family, in individuals who share the same disease-causing *RCC1* variant. Instead, we hypothesize that the nature and duration of the cellular stress also impacts the resultant phenotype, with some insults leading to greater degrees of NCT disruption, resulting in protein mislocalization, and then, after a certain threshold, causing neuronal damage. The clinical presentation indicates that the nervous system, and motor neurons in particular, are exquisitely sensitive to perturbed NCT, which perhaps reflects the lack of regenerative capacity in this tissue.

All patients were healthy prior to their presentation and, during infancy, will have been exposed to a variety of infectious agents. Understanding why certain pathogens act as precipitants for this disorder, but others do not, will be of direct clinical relevance and will provide further insights into the role of NCT in health and disease. It is likely that this condition is under ascertained, as a genomic explanation is often not considered in an acute setting when a child presents after infection with a rapid neurological deterioration. Therefore, analysis of *RCC1* should be considered in all acutely unwell children where genome sequencing is undertaken as part of the diagnostic work up.

Our converging data indicate that deleterious biallelic *RCC1* variants predispose neurons to a sudden and profound disruption of the Ran-GTP gradient, impairing normal NCT, and leading to mislocalization of proteins and mRNA. Loss of the Ran-GTP gradient and NCT disruption have been observed in both sporadic and familial cases of ALS, though this dysfunction occurs over a longer time, resulting in progressive deterioration over several years.^7,27,28^ The clinical presentation, early onset and brain involvement, seen in this cohort are not typical of ALS, but the cytoplasmic aggregation of TDP-43 and predilection for motor axons suggest that the underlying cellular mechanisms in both disorders converge around NCT dysfunction. Correcting impaired NCT is considered an attractive therapeutic target in ALS, but only a small number of drugs have been developed which modify nuclear transport.^10,11,29^ The findings from this cohort, and other NCT disorders, may prove valuable when developing novel, effective treatments, which may in-turn inform the management and symptomatic prevention of families affected by this disorder.

## Data Availability

All data produced in the present study are available upon reasonable request to the authors

https://www.genomicsengland.co.uk/research

## Acknowledgments

We acknowledge grant support from: the Wellcome Trust (PhD studentship to JRH); the Manchester NIHR BRC (NIHR203308); LifeArc Pathfinder award; and NIHR (JHM is a NIHR Doctoral Fellow, 301748). This research was made possible through access to data in the National Genomic Research Library, which is managed by Genomics England Limited (a wholly owned company of the Department of Health and Social Care). The National Genomic Research Library holds data provided by patients and collected by the NHS as part of their care and data collected as part of their participation in research. The National Genomic Research Library is funded by the National Institute for Health Research and NHS England. The Wellcome Trust, Cancer Research UK and the Medical Research Council have also funded research infrastructure. Further acknowledgements of funding are provided in the supplementary appendix.

## Data Accessibility Statement

The variants were submitted to Clinvar with the following accession numbers SCV004231856 - SCV004231862 and SUB14705452. Research on the de-identified patient data from the 100,000 Genomes Project used in this publication can be carried out in the Genomics England Research Environment subject to a collaborative agreement that adheres to patient led governance. All interested readers will be able to access the data in the same manner that the authors accessed the data. For more information about accessing the data, interested readers may contact or access the relevant information on the Genomics England website: https://www.genomicsengland.co.uk/research.

