## Supplementary Data for "Genetic Risk of Axonal Neuropathy Following Infection"

**Supplementary Information: Genetic Risk of Axonal Neuropathy Following Infection**

**Contents**


**1. Supplementary Clinical Information**

1.1 Table S1. Clinical features of patients with recessive *RCC1* variants

**2. Supplementary Methods**

2.1 Cloning of constructs for *RCC1* mammalian expression

2.2 Generation of constructs for Rcc1 and Ran protein expression

2.3 Table S2. Primers used in cloning of constructs

2.4 Thermal Shift Assay

2.5 Guanine Exchange Factor Assay

2.6 Cell culture and stress conditions

2.7 Immunofluorescence

2.8 Western Blotting

2.9 *Drosophila* studies

2.10 Table S3-5. Oligonucleotide sequences used during *Drosophila* studies

**3. Supplementary Results**

3.1 Figure S1: Patient brain magnetic resonance imaging

3.2 Table S6: *RCC1* variants identified in patients

3.3 Figure S2: Structural modelling of Rcc1 protein variants

3.4 Figure S3: Recombinant expression of Rcc1 mutant proteins.

3.5 Figure S4: Thermal stability assay response curves

3.6 Figure S5: Western blotting of Rcc1 in patient fibroblasts

**4. Supplementary References**

**5. Supplementary Acknowledgements and Institutional Support**

### Table S1. Clinical features of patients with biallelic *RCC1* variants

| | Family 1 | | | | Family 2 | | | | Family 3 | | | |
|---|---|---|---|---|---|---|---|---|---|---|---|---|
| | Patient 1 | Patient 2 | Patient 3 | Patient 4 | Patient 5 | Patient 6 | Patient 7 | Patient 8 | Patient 9 | Patient 10 | Patient 11 | Patient 12 |
| **General Information** | | | | | | | | | | | | |
| Alive/ Deceased | Alive | Deceased | Deceased | Deceased | Deceased | Deceased | Deceased | Alive | Alive | Deceased | Deceased | Deceased |
| Sex | Female | Male | Male | Female | Female | Male | Male | Female | Male | Female | Female | Male |
| Ethnicity | South Asian | South Asian | South Asian | South Asian | South Asian | South Asian | South Asian | South Asian | White European | White European | White European | White European |
| **Genetic Information** | | | | | | | | | | | | |
| Variants* (protein alteration) | c.127G>A p.(Gly43Ser) | c.127G>A p.(Gly43Ser) | c.127G>A p.(Gly43Ser) | c.127G>A p.(Gly43Ser) | c.127G>A p.(Gly43Ser) | c.127G>A p.(Gly43Ser) | c.127G>A p.(Gly43Ser) | c.127G>A p.(Gly43Ser) | c.238G>A p.(Val80Met) c.1195C>T p.(Arg399Cys) | c.238G>A p.(Val80Met) | Not tested | Not tested |
| Zygosity | Homozygous | Homozygous | Homozygous | Homozygous | Homozygous | Homozygous | Homozygous | Homozygous | Compound Heterozygous | Homozygous | N/A | N/A |
| Method of variant detection | Trio WES | Sanger | Sanger | Trio WES | Sanger | Sanger | Trio WES | Sanger | Trio WGS | Trio WES | N/A | N/A |
| gnomAD allele frequency* | 0.00 | 0.00 | 0.00 | 0.00 | 0.00 | 0.00 | 0.00 | 0.00 | 0.00002969 / 0.00002231 | 0.00002969 | N/A | N/A |
| Exon of 13 | 6 | 6 | 6 | 6 | 6 | 6 | 6 | 6 | 6 and 13 | 6 | N/A | N/A |
| Parental consan-guinity | Yes | Yes | Yes | Yes | Yes | Yes | Yes | Yes | No | Yes | Yes | Yes |
| Perinatal issues | Nil | Enlarged cisterna magna on USS 11mm | Nil | Nil | Nil | Nil | Nil | Nil | Nil | Yes | Yes | Yes |
| Pre-existing development al delay | Mild motor | Nil | Nil | Nil | Mild mixed motor | Nil | Nil | Nil | Nil | Nil | Nil | Nil |
| Medical history | Nil | Nil | Nil | Nil | Nil | Nil | Nil | Neonatal jaundice | Single right kidney | Nil | Nil | Epilepsy |
| **Presentation** | | | | | | | | | | | | |
| Age at presentation | 0-5years | 0-5 years | 0-5 years | 0-5 years | 0-5 years | 0-5 years | 0-5 years | 0-5 years | 0-5 years | 0-5 years | 0-5 years | 0-5 years |
| Preceding illness | Yes | Yes | Yes | Yes | Yes | Yes | Yes | Yes | Yes | Yes | Yes | Unknown |
| Type of illness | URTI | Diarrhea & fever | Fever & poor feeding | Roseola infantum | Fever of unknown origin | Sepsis of unknown origin | Sepsis of unknown origin | LRTI | Viral gastro-enteritis | URTI | LRTI | N/A |
| Hypotonia | Yes | Yes | Yes | Yes | Yes | Yes | Yes | Yes | Yes | Yes | Yes | NC |
| Weakness (MRC) | Yes (0/5) | Yes (0/5) | Yes (0/5) | Yes (0/5) | Yes (NC) | Yes (NC) | Yes (NC) | Yes (NC) | Yes (3/5) | Yes (0/5) | Yes (NC) | Yes (NC) |
| Reflexes | Absent | Absent | Absent | Absent | Absent | Absent | Absent | Unaffected | Absent | Absent | Absent | NC |

| | | | | | | | | | | | | |
|---|---|---|---|---|---|---|---|---|---|---|---|---|
| Bulbar involvement | Yes | Yes | Yes | Yes | Yes | Yes | Yes | No | Yes | Yes | Yes | Yes |
| Recovery | Limited | No | No | No | Limited | Limited | No | No | Complete initially | Limited | Limited | No |
| Recurrent episodes | No | No | No | No | Yes | Yes | No | No | Yes | Yes | Yes | Yes |
| GBS Considered as differential | Yes | Yes | NC | Yes | Yes | NC | NC | NC | Yes | Yes | Yes | No |
| Age at death | N/A | 0-5 years | 0-5 years | 0-5 years | 0-5 years | 10-15 years | 0-5 years | N/A | N/A | 20-25 years | 5-10 years | 0-5 years |
| **Investigations** | | | | | | | | | | | | |
| Lumbar puncture | Mildly increased protein | Mildly increased protein | NAD | NAD | Mildly increased protein | NC | NC | NAD | NC | Mildly increased protein | NAD | NC |
| EEG | Non-specific abnormal background activity | N/A | N/A | N/A | NAD | N/A | N/A | N/A | NC | NAD | NC | NC |
| Nerve conduction studies | Sensorimotor poly-neuropathy with axonal features | No motor or sensory responses | N/A | N/A | Sensorimotor poly-neuropathy | N/A | Sensorimotor poly-neuropathy | N/A | Axonal length dependent motor poly-neuropathy | Axonal motor neuropathy or neuronopathy | NC | NC |
| Brain imaging | Progressive cerebral atrophy | Progressive cerebral and cerebellar atrophy | Generalized cerebral atrophy | N/A | Progressive cerebral and cerebellar atrophy | NC | Marked atrophy with fronto-temporal predominance | N/A | NAD | Progressive cerebral and cerebellar atrophy | NA | NA |
| Virology | NAD | EBV serology positive <8 weeks prior to presentation | NAD | NAD | NAD | NAD | NAD | NAD | NAD | NAD | NC | NC |
| Muscle biopsy | Neurogenic atrophy with group atrophy pattern | N/A | N/A | N/A | Complex 1 deficiency mitochondrial chain defect | N/A | NAD | N/A | Denervation-reinnervation changes | NA | NA | NA |
| Autopsy findings | N/A | Widespread micro vacuolation of the neuropil and neuronal depletion, gliosis, axonal swelling | N/A | N/A | NC | NC | NC | NC | N/A | N/A | N/A | N/A |

| | **Family 4** | **Family 5** | | **Family 6** | **Family 7** | **Family 8** | **Family 9** | | **Family 10** | **Family 11** | **Family 12** | | **Summary** |
|---|---|---|---|---|---|---|---|---|---|---|---|---|---|
| | **Patient 13** | **Patient 14** | **Patient 15** | **Patient 16** | **Patient 17** | **Patient 18** | **Patient 19** | **Patient 20** | **Patient 21** | **Patient 22** | **Patient 23** | **Patient 24** | |
| **General Information** | | | | | | | | | | | | | |
| Alive/ Deceased | Alive | Alive | Deceased | Deceased | Alive | Deceased | Alive | Deceased | Alive | Alive | Alive | Deceased | N/A |
| Sex | Female | Female | Female | Male | Male | Male | Male | Female | Female | Female | Female | Male | 10 Male<br>12 Female |
| Ethnicity | Middle Eastern | Middle Eastern | Middle Eastern | White European | White European | Middle Eastern | Middle Eastern | Middle Eastern | White European | Middle Eastern | South Asian | South Asian | N/A |
| **Genetic Information** | | | | | | | | | | | | | |
| Variant* (protein alteration) | c.280A>G<br>p.(Asn94Asp) | c.330G>C<br>p.(Met110Ile) | Not tested | c.1195C>T<br>p.(Arg399Cys) | c.330G>C<br>p.(Met110Ile)<br>c.604G>A<br>p.(Gly202Ser) | c.330G>C<br>p.(Met110Ile) | c.330G>C<br>p.(Met110Ile) | Not tested | c.209T>C<br>p.(Val70Ala)<br>c.330G>C<br>p.(Met110Ile) | c.781G>A;<br>p.(Val261Met) | c.781G>A;<br>p.(Val261Met) | Not tested | N/A |
| Zygosity | Homozygous | Homozygous | N/A | Homozygous | Compound heterozygous | Homozygous | Homozygous | N/A | Compound Heterozygous | Homozygous | Homozygous | N/A | N/A |
| Method of variant detection | Trio WES | Singleton WES | N/A | Trio WES & Singleton WGS | Trio WES | Trio WES | Trio WES | N/A | Sanger | WES | WGS | N/A | N/A |
| gnomAD allele frequency* | 0.00 | 0.00001735 | N/A | 0.00002231 | 0.00001735 / 0.000001239 | 0.00001735 | 0.00001735 | N/A | 0.00 / 0.00001735 | 0.000003718 | 0.000003718 | N/A | N/A |
| Exon of 13 | 7 | 7 | N/A | 13 | 7 and 9 | 7 | 7 | N/A | 6 and 7 | 10 | 10 | N/A | N/A |
| Parental consan-guinity | Yes | Yes | Yes | No | No | Yes | Yes | Yes | No | Yes | Yes | Yes | N/A |
| Perinatal issues | Nil | Nil | Nil | Respiratory distress in newborn period | Nil | Nil | Nil | Nil | Nil | Nil | Nil | Oligohydramnios | 2/22 |
| Pre-existing developmental delay | Nil | Nil | Nil | Nil | Nil | Nil | Nil | Nil | Nil | Nil | Nil | Mild global | 2/22 |
| Medical history | Nil | Nil | Nil | Nil | Nil | High-arched palate | Nil | Nil | Nil | Nil | Nil | Nil | 4/22 |
| **Presentation** | | | | | | | | | | | | | |
| Age at presentation | 10-15 years | 0-5 years | 0-5 years | 0-5 years | 0-5 years | 0-5 years | 0-5 years | Unknown | 0-5 years | 0-5 years | 0-5 years | 0-5 years | median 18 months |
| Preceding illness | Nil | Yes | Yes | Yes | Yes | Yes | Yes | Yes | Yes | Yes | Yes | Yes | 22/24 |
| Type of illness | N/A | URTI | URTI | URTI & fever | Febrile infection | Influenza | NC | NC | URTI | Ear Infection | Fever following MMR vaccination | Chickenpox | N/A |
| Hypotonia | No | Yes | Yes | Yes | Yes | Yes | Yes | Yes | Yes | Yes | Yes | Yes | 24/24 |

| | | | | | | | | | | | | | |
|---|---|---|---|---|---|---|---|---|---|---|---|---|---|
| Weakness (MRC) | Yes (NC) | Yes (3/5) | Yes (NC) | Yes (NC) | Yes | Yes (3/5) | Yes (NC) | Yes (NC) | Yes | Yes | Yes | Yes (NC) | 24/24 |
| Reflexes | Unaffected | Absent | Absent | Absent | Absent | Absent | Absent | NC | Absent | Absent | Unaffected | NC | 20/24 |
| Bulbar involvement | No | Yes | NC | Yes | Yes | No | No | NC | Yes | NC | Yes | Yes | 16/24 |
| Recovery | No | No | Limited | Limited | Limited | Limited | Partial | No | Partial | Limited | Limited | No | 12/24 |
| Recurrent episodes | Progressive | Yes | Yes | Yes | Yes | Yes | No | No | Yes | Yes | Yes | No | 15/24 |
| GBS Considered as differential | NC | NC | NC | CIDP | NC | Yes, then CIDP | Yes | Yes | NC | Yes | NC | NC | 10/24 |
| Age at death | N/A | N/A | 5-10 years | 0-5 years | N/A | 10-15 years | N/A | 0-5 years | N/A | N/A | N/A | 0-5 years | 11 months – 23 years |
| **Investigations** | | | | | | | | | | | | | |
| Lumbar puncture | N/A | NAD | NC | Raised protein during acute episodes | NAD | NC | NC | NC | Raised protein during acute episodes | Yes | TBC | Normal | N/A |
| EEG | N/A | N/A | N/A | Abnormal background activity | Poor basal activity | NAD | NC | NC | NC | NC | TBC | NC | N/A |
| Nerve conduction studies | Motor Neuropathy | Axonal motor neuropathy | Axonal motor neuropathy | Severe combined neuropathy | Axonal sensorimotor poly-neuropathy | Sensorimotor poly-neuropathy | Axonal sensorimotor poly-neuropathy | NC | Sensory motor neuropathy with axonal type | Sensory motor neuropathy with axonal type | Sensory Motor Axonal Neuronopathy | Mixed axonal and demyelinating sensorimotor neuropathy | N/A |
| Brain imaging | NAD | Fronto-temporal cerebral atrophy | NAD | Progressive cerebellar and cerebral atrophy | Progressive cerebral atrophy with fronto-temporal predominance | NC | NC | NC | Normal | Normal | TBC | NC | N/A |
| Virology | NAD | NAD | NC | NAD | NAD | NAD | NAD | NC | NC | NC | TBC | N/A | N/A |
| Muscle biopsy | N/A | N/A | NC | Atrophy with signs of neurogenic damage | Atrophy with secondary neurogenic changes | N/A | Lipid increase in muscle fibers | NC | NC | N/A | TBC | N/A | N/A |
| Autopsy findings | N/A | N/A | NC | N/A | N/A | NC | N/A | NC | N/A | N/A | N/A | N/A | N/A |

**Footnote:** N/A = Not Applicable. NC = Not Communicated or data unavailable. NAD = No abnormalities detected. MRC = Medical Research Council Scale.

#### 2. Supplementary Methods

##### 2.1 Cloning of constructs for *RCC1* mammalian expression

To generate the *RCC1* mammalian expression vectors, pcDNA3.1 (Thermofisher) was digested with KpnI and EcoRI. HiFi assembly was used to insert mScarlet (amplified using F1F/F1R on addgene #129721) and a synthesised gBlock fragment (IDT) containing the ORF for RCC1 (NM_001381865.2) and a Gly-Ser-Waldo linker[1]; (RCC1_gBlock), to create pCMV-mScarlet-GSW-RCC1wt (**Table S2**). This plasmid was digested with BsrGI and Bsu36I and used as the backbone to make the following constructs. c.127G>A, Gly43Ser, c.238G>A, Val80Met, c.280A>G, Asn94Asp, c.330G>C, p.Met110Ile, using a-d_F1F/a-d_F2R and mutant specific primers to introduce the mutation (**Table S2**). The plasmid was also digested with Bsu36I and EcoRV to make the following constructs c.604G>A, Gly202Ser, c.767C>T, Ser256Phe, c.1195C>T, and Arg399Cys, using e-g_F1F/e-g_F2R (**Table S2**) and mutant specific primers to introduce the mutation; all were generated via a two fragment HiFi assembly.

Introduction of c.209T>C, Val70Ala and c.781G>A, Val261Met variants into the pCMV-mScarlet-GSW-RCC1wt sequence was then performed using Q5 site-directed mutagenesis (New England Bioscience) using primer pairs j_V70A_F/R and k_V261M_F/R respectively (**Table S2**). All sequences were confirmed by DNA sequencing.

##### 2.2 Generation of constructs for Rcc1 and Ran protein expression

To conduct in vitro assays, constructs were generated for the expression of recombinant Rcc1 and Ran proteins. *RAN* cDNA was amplified from control cDNA and *RCC1* cDNA was amplified or from mammalian expression vectors containing relevant variants (**Section 2.3**) using primers designed to introduce 3’ His-tags (primers h_His_Rcc1F/R and h_His_RanF/R). His-tagged *RAN* and *RCC1* c.DNA was cloned into low copy pET-28a plasmid suitable for protein expression using 2 fragment HiFi assembly. Introduction of c.209T>C, Val70Ala and c.781G>A, Val261Met variants into the *RCC1* WT sequence was then performed via Q5 site-directed mutagenesis (New England Bioscience) using primer pairs j_V70A_F/R and k_V261M_F/R respectively (**Table S2**). All variants were confirmed by DNA sequencing.

Protein was expressed in E. *coli* Rosetta2 (DE3) bacteria (Novagen) in 500ml Overnight Express media (Novagen) for 36 hours at 37 °C. Cultures were pelleted by centrifugation at 7000 rpm for 15 min at 4 °C, and cells lysed in lysis/wash buffer (20 mM Tris-Cl pH 7.4, 150 mM NaCl, 0.1 M DTT, 0.02 % Tween-20, 10 mM imidazole, 15 % glycerol). Cell lysis was completed with 3x sonication for 10 sec at 30 % amplitude. Lysate was cleared by centrifugation at 17000 rpm for 15 min at 4 °C. His-tagged proteins were column-purified using His-Select Nickel Affinity Gel (Sigma). Purified proteins were eluted in 4x 1 mL fractions with elution buffer (20 mM Tris-Cl pH 7.4, 150 mM NaCl, 0.1 M DTT, 0.02 % Tween-20, 250 mM imidazole, 15 % glycerol), and purification was evaluated via SDS-PAGE (**Figure S2**). Once purity was confirmed protein fractions were dialyzed in dialysis buffer (20 mM Tris-Cl pH 7.4, 150 mM NaCl, 0.1 M DTT, 15 % glycerol) overnight at 4 °C to facilitate removal of imidazole. Proteins were aliquoted and stored at -80 °C until use.

#### 2.3 Table S2. Primers used in cloning of vectors

Table S2. Primers used in cloning and mutagenesis.

| Name | Sequence |
|---|---|
| RCC1_gBlock | CGGCATGGACGAGCTGTACAAGGGTGGATCCGGCGGAAGCGGCTC<br>CGCTGGCTCTGCTGCCGGATCTGGCGAGTTCatgtcacccaagcgcatagctaaaa<br>gaaggtcccccccagcagatgccatccccaaaagcaagaaggtgaaggtctcacacaggtcccacagcaca<br>gaacccggcttggtgctgacactaggccagggcgacgtgggccagctggggctgggtgagaatgtgatgga<br>gaggaagaagccggccctggtatccattccggaggatgttgtgcaggctgaggctgggggcatgcacaccgt<br>gtgtctaagcaaaagtggccaggtctattccttcggctgcaatgatgagggtgccctgggaagggacacatcag<br>tggagggctcggagatggtccctgggaaagtggagctgcaagagaaggtggtacaggtgtcagcaggagac<br>agtcacacagcagccctcaccgatgatggccgtgtcttcctctggggctccttccgggacaataacggtgtgatt<br>ggactgttggagcccatgaagaagagcatggtgcctgtgcaggtgcagctggatgtgcctgtggtaaaggtgg<br>cctcaggaaacgaccacttggtgatgctgacagctgatggtgacctctacaccttgggctgcggggaacaggg<br>ccagctaggccgtgtgcctgagttatttgccaaccgtggtggccggcaaggcctcgaacgactcctggtcccca<br>agtgtgtgatgctgaaatccaggggaagccggggccacgtgagattccaggatgccttttgtggtgcctatttca<br>cctttgccatctcccatgagggccacgtgtacggcttcggcctctccaactaccatcagcttggaactccgggca<br>cagaatcttgcttcataccccagaacctaacatccttcaagaattccaccaagtcctgggtgggcttctctggtgg<br>ccagcaccatacagtctgcatggattcggaaggaaaagcatacagcctgggccgggctgagtatgggcggct<br>gggccttggagagggtgctgaggagaagagcatacccaccctcatctccaggctgcctgctgtctcctcggtg<br>gcttgtggggcctctgtggggtatgctgtgaccaaggatggtcgtgttttcgcctggggcatgggcaccaactac<br>cagctgggcacagggcaggatgaggacgcctggagccctgtggagatgatgggcaaacagctggagaacc<br>gtgtggtcttatctgtgtccagcgggggccagcatacagtcttattagtcaaggacaaagaacagagctgaGA<br>ATTCTGCAGATATCCAGCACAGTGG |
| a-d_F1F | GCGGCATGGACGAGCT |
| a-d_F2R | catcaccaagtggtcgtttcc |
| a_G43S_F1R | cgtcgcTctggcctagtgtcag |
| a_G43S_F2F | cactaggccagAgcgacgtgggccag |
| b_V80M_F1R | tagacacaTggtgtgcatgcccccag |
| b_V80M_F2F | ggcatgcacaccAtgtgtctaagcaaaagtggcc |
| c_VN94D_F1R | cctcatcatCgcagccgaaggaatagacc |
| c_VN94D_F2F | tccttcggctgcGatgatgagggtgccctgg |
| d_M110I_F1R | ccagggacGatctccgagccctccac |
| d_M110I_F2F | gctcggagatCgtccctgggaaagtggagc |
| e-f_F1F | gcctgtggtaaaggtggcc |
| e-f_F2R | CCGCCACTGTGCTGGAT |
| e_S256F_F1R | gccctcatggAagatggcaaaggtgaaataggc |
| e_S256F_F2F | cctttgccatctTccatgagggccacgtgtac |
| f_R399C_F1R | gataagaccacacAgttctccagctgtttgccc |
| f_R399C_F2F | cagctggagaacTgtgtggtcttatctgtgtccag |
| g_G202S_F1R | ggcctagctggcTctgttccccgcagccc |
| g_G202S_F2F | gaacagAgccagctaggccgtgtgc |

| | |
|---|---|
| h_His_Rcc1F | GGGGGGATCCCATATGTCACCCAAGCGCATAGCTAAAAGAAGG |
| h_His_Rcc1R | GGGGGATCCTCAGCTCTGTTCTTTGTCCTTGAC |
| h_His_RanF | GCCGGGATCCCATATGGCTGCGCAGGGAGAGCCCCAGG |
| h_His_RanR | GCGCGGATCCTCACAGGTCATCATCCTCATCC |
| j_V70A_F | GGAGGATGTTgcgCAGGCTGAGG |
| j_V70A_R | GGAATGGATACCAGGGCC |
| k_V261M_F | TGAGGGCCACatgTACGGCTTCG |
| k_V261M_R | TGGGAGATGGCAAAGGTGAAATAG |

#### 2.4 Thermal Stability Assay

To assess thermal stability of recombinant Rcc1 proteins, thermal stability assays were conducted using Protein Thermal Shift™ Dye Kit (Applied Biosystems). 0.3 µg recombinant Rcc1WT or Rcc1G43S protein was combined with Protein Thermal Shift™ Dye Kit Buffer and 8x Dye in 20 µL reactions in a 96 well PCR plate. Reactions were also conducted with the addition of 0.3 µg recombinant Ran protein. ROX reporter dye fluorescence was measured using a StepOnePlus™ Real-Time PCR System (Applied Biosystems) after each 1 % increase in temperature between 25 °C and 99 °C. Melting temperature (TM) was derived from the peak in fluorescence from plotting melt curves of fluorescence against temperature. Reactions were conducted in triplicate within each assay. Data represent the mean of three assays ± standard deviation.

#### 2.5 Guanine Exchange Factor Assay

To establish the ability of recombinant Rcc1 to facilitate guanine exchange with Ran GTPase, GEF assays were performed using the GTPaseGlo™ Assay kit (Promega). Addition of GEF buffer containing $Mg^{2+}$ ensures nucleotide loading is catalyzed solely by the GEF (Rcc1). Assays were conducted with 7.5 µg Ran protein, combined with 5 µg $Rcc1^{WT}$ or $Rcc1^{G43S}$ in 10 μL reactions with 10 μM GTP and 1 mM DTT. GTPase reactions were incubated for four hours at 37 °C. Residual GTP was converted to ATP by addition of 10 μL GTPase-Glo Reagent and incubation at room temperature with agitation for 30 min. Bioluminescence was generated from converted ATP by addition of 20 μl Ultra-Glo™ Recombinant Luciferase Detection Reagent. Luminescence was read using Infinite 200 PRO microplate reader (Tecan) 20 min after addition of detection reagent. Luminescence values were normalized to reactions without Ran or Rcc1. Reactions were conducted in triplicate wells within each assay and data represent mean of three assays ± standard deviation.

**2.6 Cell culture and stress conditions**

**2.6.1 Fibroblasts**

Patient or control-derived primary dermal fibroblasts cells were maintained under standard culture conditions (37.0°C, 5.0% $CO_2$) in Dulbecco's Modified Eagle medium (DMEM; Sigma) with 10% foetal bovine serum (FBS; Sigma) and 1% penicillin-streptomycin (Sigma). To assess the effect of exogenous stress on cell activity, fibroblasts were subject either to (i) heat shock by placing in an incubator set to 43.0°C, 5.0% $CO_2$ for 4-20 hours or (ii) oxidative stress by incubation with DMEM containing 200μM or 500μM $H_2O_2$ for one hour while maintained under standard culture conditions. Cells were then fixed with 3.7% formaldehyde for immunofluorescence or harvested with 1x trypsin (Gibco), pelleted and snap frozen using dry ice and stored at -80°C for later protein analysis.

**2.6.2 tsBN2 cells**

The thermosensitive tsBN2 hamster cell line[2] was obtained from the RIKEN Bioresource Institute, Japan. Cells were maintained under low temperature conditions as directed (32.0°C, 5.0% $CO_2$) in Dulbecco's Modified Eagle medium (DMEM; Sigma) with 10% foetal bovine serum (FBS; Sigma) and 1% penicillin-streptomycin (Sigma). To assess the effect of exogenous stress on cell activity, tsBN2 cells were subject either to (i) heat shock by placing in an incubator set to 39.0 °C, 5.0% $CO_2$ for 4-20 hours or (ii) oxidative stress by incubation with DMEM containing 200µM or 500µM $H_2O_2$ for one hour while maintained under standard culture conditions. Cells were then fixed with 3.7% formaldehyde for immunofluorescence or harvested with 1x trypsin (Gibco), pelleted and snap frozen using dry ice and stored at -80°C for later protein analysis.

2.6.3 Transfection of Mammalian cell lines

The tsBN2 cell line was transfected with the pCMV-mScarlet-GSW-RCC1 vectors containing variants identified in affected individuals. Prior to transfection, 30,000 cells were seeded 11 mm round glass coverslips in 24 well plates. Cells were then transfected with 200ng purified vector using 1µl Lipofectamine™ 2000 (Invitrogen) per well. Cells were cultured overnight before induction of stress conditions and fixing for immunofluorescence. Only mScarlet$^{+}$ cells were used during downstream quantification.

#### 2.7 Immunofluorescence

To assess Rcc1 localisation and response to heat shock, immunofluorescence of fibroblasts and tsBN2 cells was conducted. 30,000 cells were seeded on sterile glass coverslips in 24 well plates. After 24 hours cells were subject to stress conditions as outlined in 2.8.1-2. Cells were washed twice with warm phosphate buffered saline (PBS, Gibco), fixed with 3.7% formaldehyde (Sigma) for 15 min at room temperature, then permeabilized with 0.1 % Triton-X 100 solution (Sigma). Cells were washed twice in PBS and were blocked with 1.0 % bovine serum albumin (BSA, Sigma) for 30 min at room temperature. Cells were incubated primary antibodies: anti-Rcc1 rabbit polyclonal antibody (22142-1-AP, Proteintech), anti-Ran mouse monoclonal antibody (610341, BD Biosciences), anti-TDP-43 rabbit polyclonal antibody

(10782-2-AP, Proteintech), or anti-Phospho-TDP43 (Ser409/410) mouse monoclonal antibody (66318-1-Ig, Proteintech) overnight at 4°C. Cells were washed twice with PBS before applying secondary Alexafluor-488 Goat anti-Rabbit (AB_2338046, Jackson ImmunoResearch) and AlexaFluor-647 Goat anti-Mouse (AB_2535804, Invitrogen) in 1.0 % BSA for 1 hour at room temperature in the dark. To counterstain nuclei, coverslips were incubated with 300nM DAPI for 15 min at room temperature. Cover slips were washed twice with PBS, then adhered to glass microscope slides with ProLong Diamond Anti-fade Mountant (Invitrogen) overnight at room temperature in the dark. Slides were imaged within one week of staining using a Zeiss Axio Imager.D2 upright microscope using a 20x, 40x or 63x objective and captured using a Coolsnap HQ2 camera (Photometrics) through Micromanager software v1.4.23. Images were processed and analyzed using Fiji ImageJ (http://imagej.net/Fiji/Downloads).

##### 2.8 Protein Extraction and Western blotting

Protein lysates were extracted by whole cell lysis, or by extracting soluble and insoluble fractions. Whole cell protein lysates were harvested from cell pellets thawed on ice by resuspending in 200µl RIPA buffer (Sigma), followed by homogenization by shaking on ice for 15 min, before disrupting DNA with 25G needle and 1ml syringe. Lysates were cleared by centrifugation at 17,000x g for 15 min.

Soluble and insoluble fractions were harvested by resuspending cells in 200µl RIPA buffer and homogenization on ice for 15 min. Lysates were centrifuged to pellet insoluble material. Supernatants were removed to separate tubes and RIPA-insoluble pellets were resuspended in 7M urea buffer (7 M urea, 2 M thiourea, and 4% (w/v) CHAPS, in 30 mM Tris–HCl (pH 8.5)) and homogenised with 25G needle and 1ml syringe and incubated while shaking at room temperature for 15 min.

Lysate protein concentration was measured using a BCA assay (ThermoFisher Scientific). Sample concentrations were equalized during preparation for SDS-PAGE with 4x Lamelli buffer (Invitrogen), 10% β-mercaptoethanol and $dH_2O$, before denaturing protein by incubation at 90°C for 10 min. 10μg protein was separated via SDS-PAGE using 12% mini-PROTEAN gels (Bio-Rad) for 1 hour at 180V. Proteins were transferred to PVDF membranes using a Trans-Blot Turbo Transfer system (Bio-Rad) at 1.3 A constant for 11 min. Membranes were blocked in 2% BSA in tris buffered saline with 0.1% weight/volume Tween-20 (TBST) for one hour. Primary antibodies anti-Rcc1 rabbit polyclonal antibody (22142-1-AP, Proteintech), anti-beta actin monoclonal antibody (99009-1-Ig, Proteintech) , anti-TDP-43 rabbit polyclonal antibody (10782-2-AP, Proteintech), or anti-Phospho-TDP43 (Ser409/410) mouse monoclonal antibody (66318-1-Ig, Proteintech) overnight at 4°C. Membranes were washed twice in TBST before incubation with secondary antibodies (IRDye 800 Goat anti-Rabbit and/or IRDye 680 Donkey anti-mouse H+L (both LicorBio) for 1 hour at room temperature. Membranes were washed 4x for 10 min in TBST before visualizing bands using a Licor OdysseyXF infrared imaging system (LicorBio). Images were quantified using Fiji ImageJ (http://imagej.net/Fiji/Downloads) by normalization to beta actin.

#### 2.9 *Drosophila* Studies

##### 2.9.1 Construct generation for fly CRISPR-Cas9

The HDR plasmid containing human RCC1 (hRCC1) coding sequence with mScarlet-I fused at the N-termini, together with *3xP3-eGFP* marker, was assembled to pUC57 digested with *Eco*RI and *Bam*HI using a HiFi assembly kit (New England Biolabs) and primers provided in **Table S3**. Four 20 bp gRNA sequences (**Table S4**) with no predicted off-targets were selected using the FlyCRISPR target finder (http://targetfinder.flycrispr.neuro.brown.edu)[3] and cloned individually into the *Bbs*I site of pU6-BbsI-chiRNA (Addgene #45946).

##### 2.9.2 Fly strains

For generation of CRISPR-Cas9 knock-ins, all vectors were midi-prepped and eluted in $ddH_2O$. gRNA plasmids (60ng/ul each) were injected with HDR plasmid (500ng/ul) into *nanos-Cas9* embryos before crossing individual surviving adults to a *v*, *cho* strain. Transgenic offspring were selected using the 3P3-GFP marker before sequencing to confirm correct insertion. For genotyping, genomic DNA was extracted from individual flies by crushing with a pipette tip in 50ul of squishing buffer (10mM Tris HCL (pH8), 1mM EDTA, 25Mm NaCl, 200µg/ml Proteinase K) and incubating the solution at 37C for 30 min before inactivating the Proteinase K by heating to 95C for 3 min. After centrifugation, supernatant was used to PCR amplify across the CRISPR-Cas9 breakpoints using primers described in **Table S5** and PCR products were sent to Genewiz for Sanger sequencing.

The genotype of fly strains used in this study are given below:

*elav-GAL4* (Richard Baines, University of Manchester)

$w^{1118}$; *UAS-RCC1*$^{IR}$ (GD6989, v38389, Vienna Drosophila Research Centre)

$y^1$ $sc^1$ $v^1$;; {$y^{+t7.7}$ $v^{+t1.8}$ *nanos-Cas9*} *attp2* (from Simon Collier, the University of Cambridge)

*v cho* and *v cho*;; *TM6B*/*MKRS* (from Levente Kovacs, University of Cambridge)

*v cho*;; {*mScarlet-hRCC1*$^{wt}$ *3xPS-GFP*}/*TM6B* (this study)

*v cho*;; {*mScarlet-hRCC1*$^{G43S}$ *3xPS-GFP*}/*TM6B* (this study)

##### 2.9.3 Lightsheet imaging

Adult fly brains were dissected in PBS, fixed in 4% formaldehyde for 30 mins and washed in PBS. Samples were mounted in 2% low gelling temperature agarose in glass capillaries before imaging on a Zeiss Z.1 Lightsheet equipped with 20x Plan-Apochromat 1.0 NA objective. Images were analyzed using Imaris (version 10.01).

##### 2.9.4 *Drosophila* survival measurements:

To induce oxidative stress, food was supplemented with Paraquat (PQ, Sigma). Fly food consisted of the following ingredients dissolved in water: 78 g/L glucose, 72 g/L maize, 50 g/L yeast, 10 g/L agar, 27 ml/L Nipagen, 3 ml/L Proprionic Acid. For chronic treatments of mScarlet-hRCC1 lines, staged 3rd instar larvae were transferred to food supplemented with 30 mM PQ and the number of animals that eclosed were counted to determine overall survival. For repeated PQ treatments, 10-day old adult male flies were treated with repeated exposures of 20 mM PQ for 24h, followed by PQ withdrawal for 48h. For more acute treatments, flies were treated with repeated exposures of 50mM PQ for 24h followed by PQ withdrawal for 24h.

**2.9.5 Measurements of *Drosophila* activity:**

The *Drosophila* Activity Monitor (DAM) system (TriKinetics, Waltham, MA) was used to assess the locomotor behavior of *Drosophila* melanogaster at 25ºC with constant humidity and day/night light cycle. This system tracks individual fly movements by recording interruptions in infrared beams as flies traverse sealed glass tubes with food at one end and cotton wool at the other.[4] Flies were allowed to acclimate to the experimental conditions for at least 24 h, after which behavioral observations were monitored. Data were analyzed using Rtivity software.[5]

#### 1.10 Tables S3-5. Oligonucleotide sequences used during *Drosophila* studies

**Table S3.** Primer sequences for CRISPR-Cas9 constructs and genotyping

| Name | Fragment to amplify | sequence |
|---|---|---|
| BN05_F1F | 5' homology arm | aaacgacggccagtgaagttaatggaaaaccttttcaaatagtattgtttattttctgat |
| BN05_F1R | | CATagtcagggcctttctgcgcggcaatttagcttgtattttcaca |
| BN05_F2F | mScarlet-human RCC1 | cagaaaggccctgactATGGTGAGCAAGGGCGAG |
| BN05_F2R | | CTAGGGttaatcagctctgttctttgtccttgactaataagac |
| BN05_F3F | 3' transposon end | cagagctgattaaCCCTAGAAAGATAATCATATTGTGACGTACGTT |

| | | |
|---|---|---|
| BN05_F3R | | CTCACCATGGTGGCGACCGGCTTCG |
| BN05_F4F | eGFP | CGCCACCATGGTGAGCAAGGGCGAG |
| BN05_F4R | | CGCGGCCGCTACTTGTACAGCTCGTCCATGCCGAG |
| BN05_F5F | 5' transposon end | TACAAGTAGCGGCCGCGACTCTAGATCAT |
| BN05_F5R | | cgcttggcttaaCCCTAGAAAGATAGTCTGCGTAAAATTGACG |
| BN05_F6F | 3' homology arm | CTAGGGttaagccaagcggggcggtaaaaag |
| BN05_F6R | | Agtcgacgggcccggagtaaatgcaacttgttcacctctgtacac |
| BN04a_G43S_F | Gly43Ser site-directed mutagenesis | cactaggccagAgcgacgtgggccagc |
| BN04a_G43S_R | Gly43Ser site-directed mutagenesis | cgtcgcTctggcctagtgtcag |

**Table S4.** Guide RNA sequences for humanised fly strains

| | | |
|---|---|---|
| gRNA1 | 5' end of fly Rcc1 ORF | ctgactaataacaacaatgc |
| gRNA2 | 5' end of fly Rcc1 ORF | cggcattgttgttattagtc |
| gRNA3 | 3' end of fly Rcc1 ORF | aagcctgctgccaagcgggg |
| gRNA4 | 3' end of fly Rcc1 ORF | Aagaagcctgctgccaagcg |

**Table S5**. Primers used for genotyping

| | | |
|---|---|---|
| BN05_gF1 | Located outside of the 5H arm | gcggaatgggcaaagtggat |
| BN05_mSacrlet_gR1 | Located at 5' end of mScarlet | TCCATGTGCACCTTGAACCG |
| BN05_transposon_gF2 | Located at 5' transposon end | CCTAAATGCACAGCGACGGA |
| BN05_gR2 | Located outside of the 3H arm | aaccctcaactgtgcatgcc |

##### 3.1 Patient Brain Magnetic Resonance Imaging

**Patient 1:**

Selected T2-weighted axial, diffusion weighted and ADC map images from the MRI Head a day following cardiac arrest demonstrated marked diffusion restriction in the splenium of the corpus callosum, with subtle associated high signal on T2 images (arrows), and subtle diffusion restriction in the frontoparietal white matter (corona radiata), without discernible signal abnormality on T2 weighted imaging (arrowheads). These findings are most likely to represent hypoxic ischemia encephalopathy secondary to cardiac arrest.

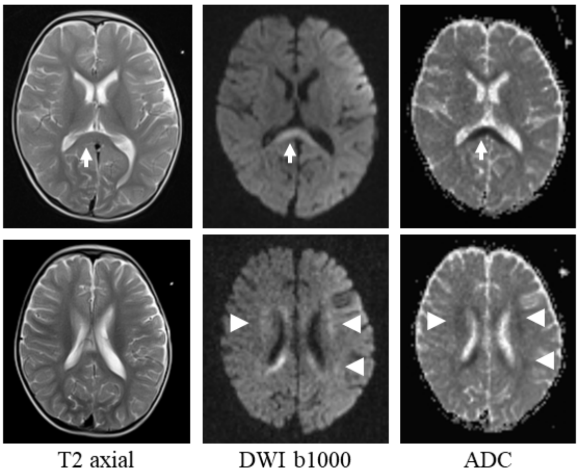


MR findings on follow up:

Selected T1 sagittal, T1 and T2 axial and FLAIR coronal images from follow-up MR scans 1.5 months and 8 months post-presentation show considerable progressive atrophy of the cerebral parenchyma, including the corpus callosum (arrows), when compared with the initial MR. There is increased prominence of the ventricles and extra-axial CSF spaces. Atrophy is global, but there is definite fronto-temporal predominance with relative sparing of the parieto-occipital regions. Progressive atrophy is beyond what would be expected following the ischemic injury at presentation, both in distribution and timeframe. No signal abnormality or diffusion restriction on the follow up imaging.

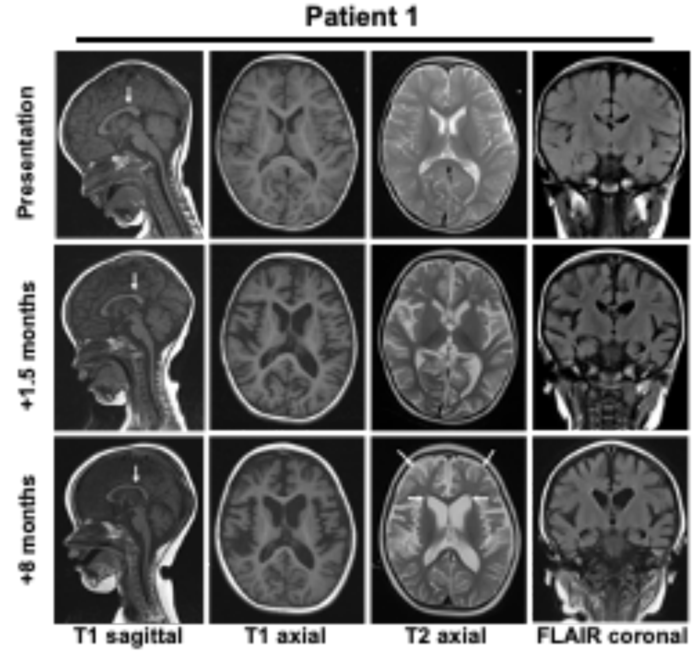

#### Patient 5

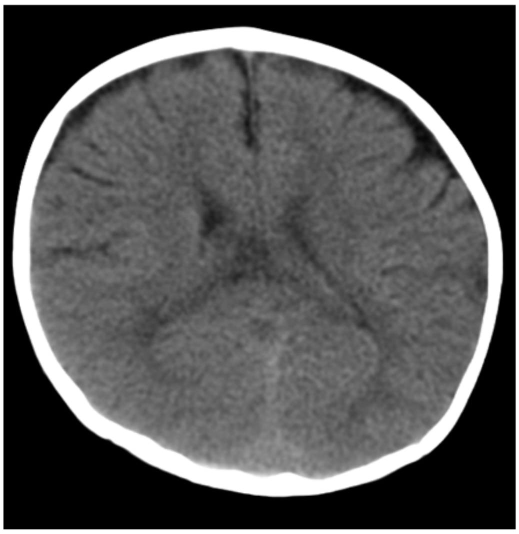

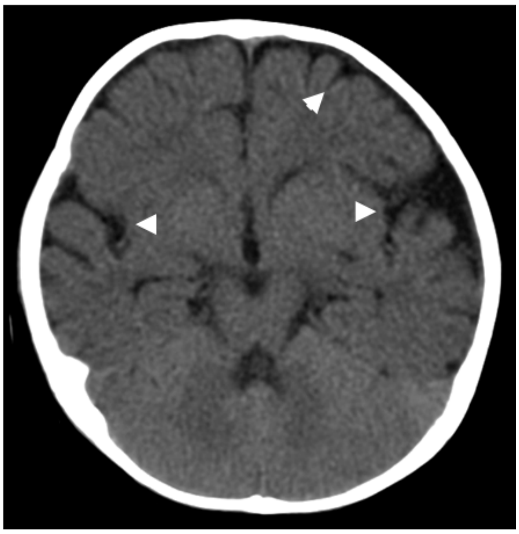

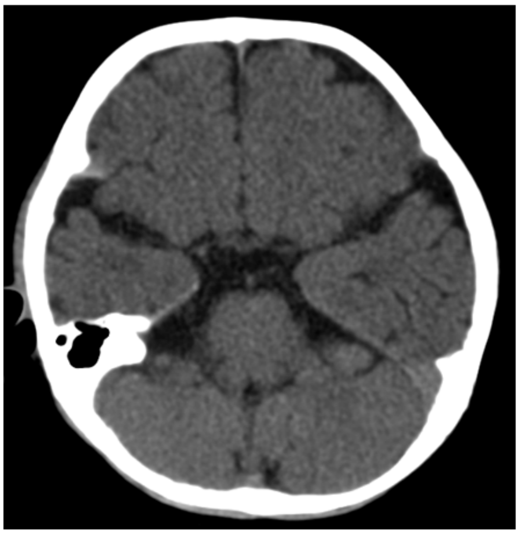

Axial CT images of the head at 9.5 months after presentation show atrophy which is predominantly fronto-temporal and slightly asymmetrical, affecting left more than right. Associated prominence of the CSF spaces including sylvian fissures (arrowheads).

Note, benign prominence of the CSF spaces, a differential diagnosis for CSF prominence in a neonate, tends to involve only the frontal poles, whereas this involves the whole convexity.

### Patient 7

**MR Findings**

T2 axial (A & B), T1 sagittal (C) and T2 coronal (D) MR images of the brain show marked atrophy, with frontal predominance (arrows) with prominence of the frontal horns of the lateral ventricles (arrowheads) and prominence of the sylvian fissures.

Deep nuclei and corpus callosum spared, as are parieto-occipital regions and cerebellum.

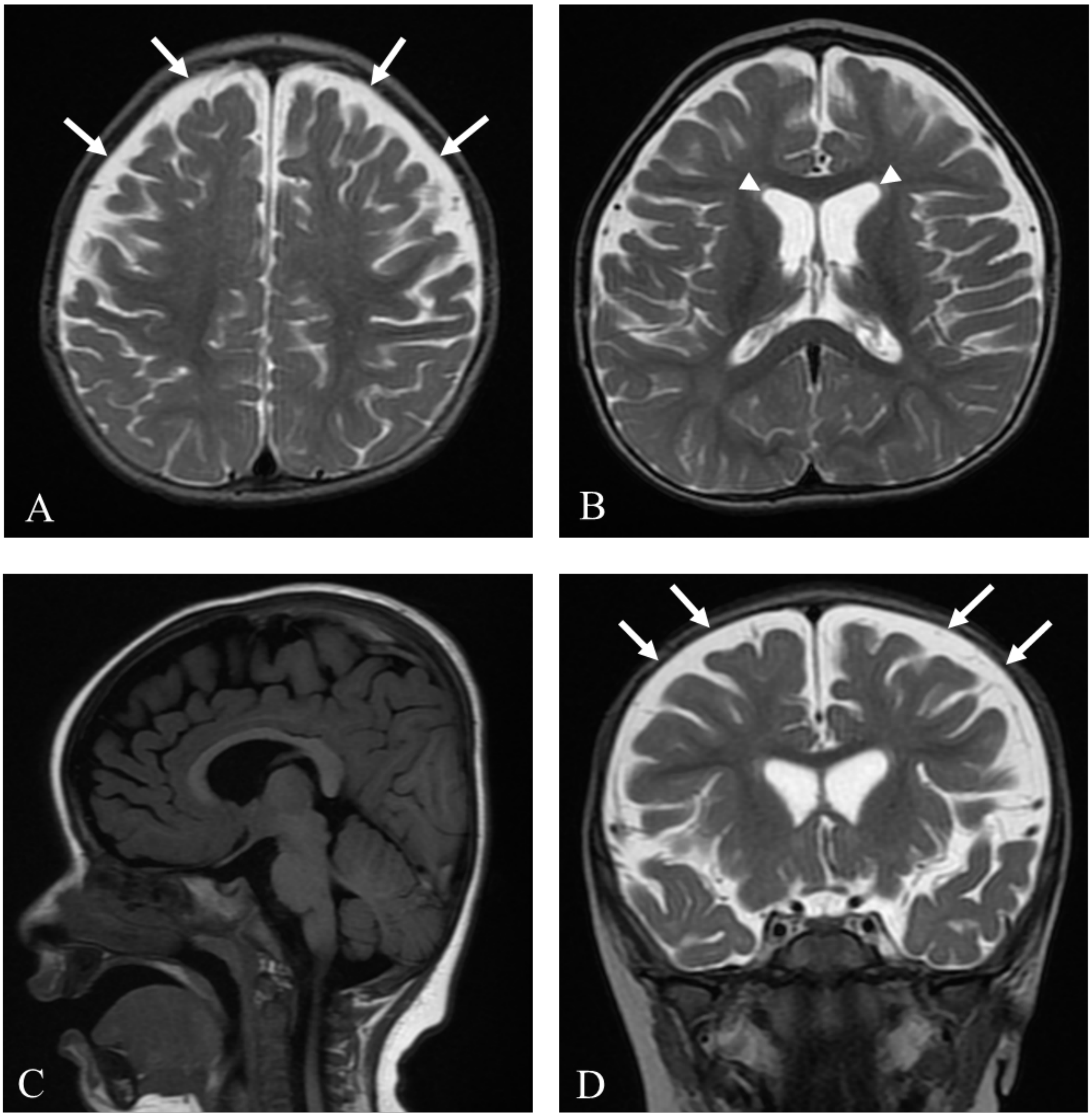

#### Patient 9

**MR Findings**

Selected T2 axial (A & B), T2 FLAIR coronal (C) and T1 sagittal (D) MR images of the brain are normal in appearance.

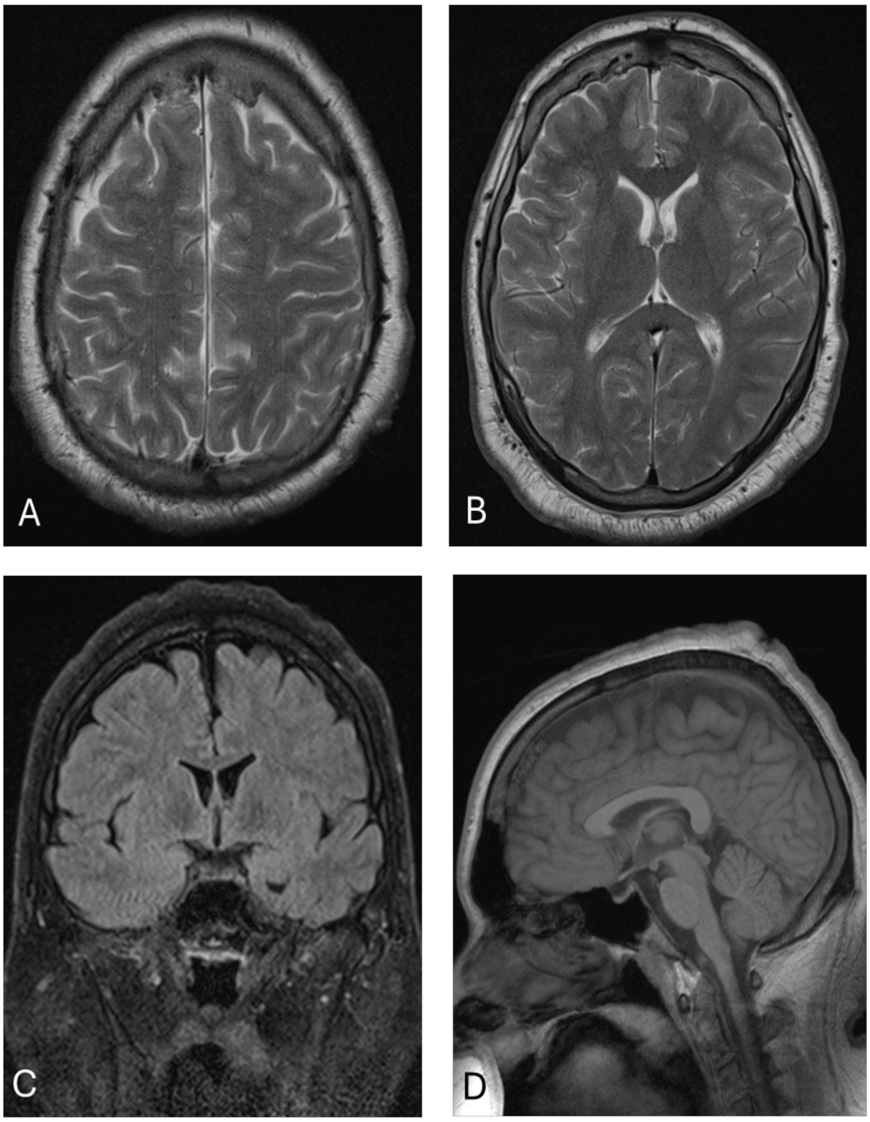

**Patient 13**
Selected T2 FLAIR axial MR images of the brain at the time of presentation (A) and 2 years after presentation (B) are normal.

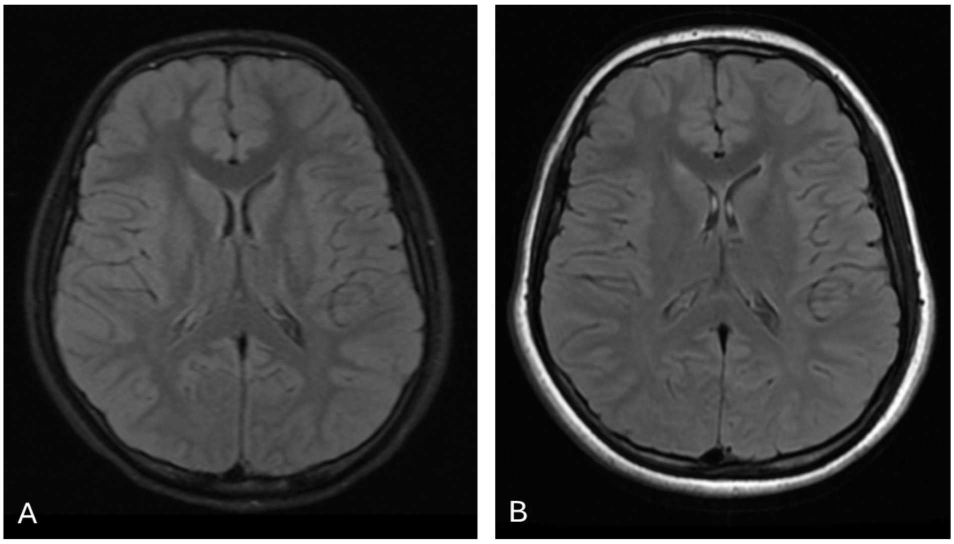


**Patient 14**
Selected T2 axial and T2 FLAIR coronal images from an MR. Head degraded by movement artefact. Mild but definite frontotemporal cerebral atrophy with associated CSF space predominance, including the ventricles.
No pathology was reported. Normal MR spine.

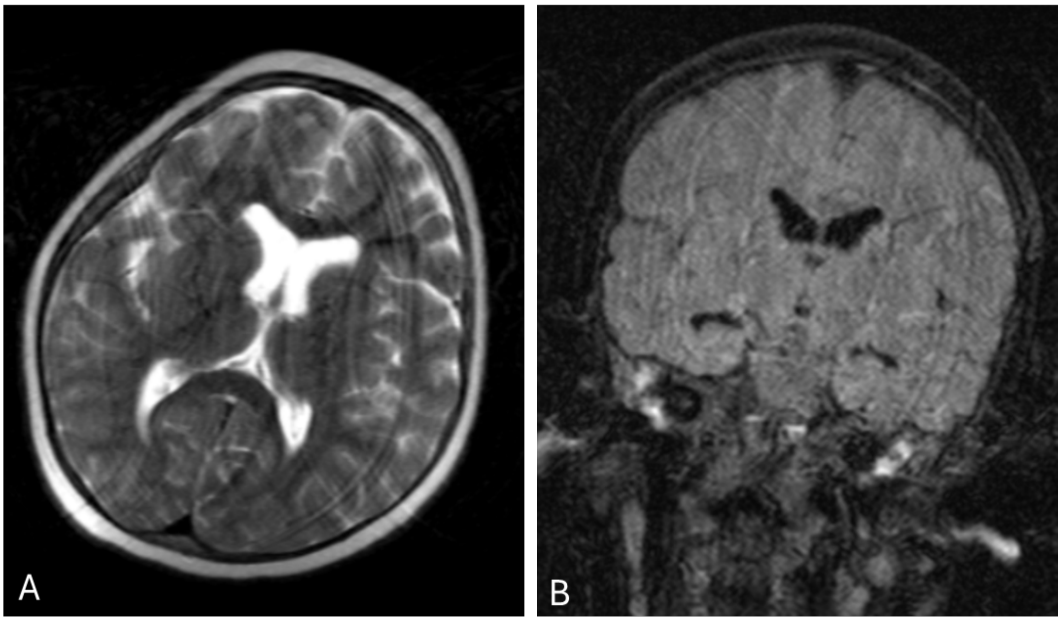

**Patient 16**

Brain findings at presentation: Symmetrical high signal changes in the periventricular and parieto-occipital white matter on selected T2 FLAIR axial (A,B) and coronal (C) imaging without enhancement or diffusion restriction (arrows).

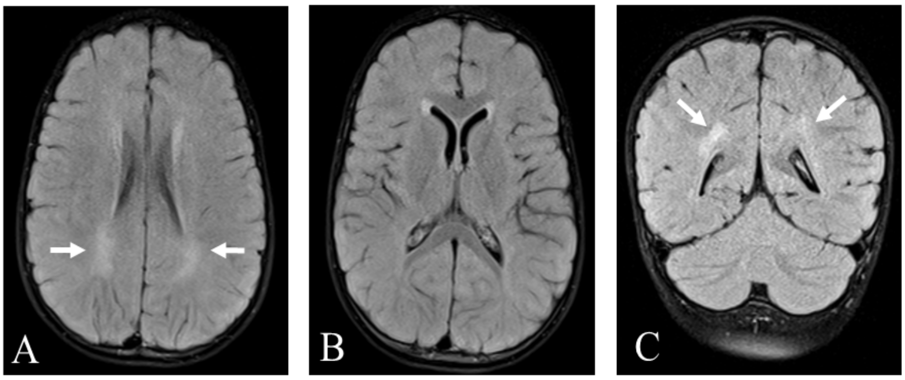


MR spine 3 months after presentation: Selected T2 sagittal (A, B) and T1 sagittal (C) images of the spine demonstrate subtle high T2 signal in the cord, particularly in the dorsal columns of the cervical cord (arrows).

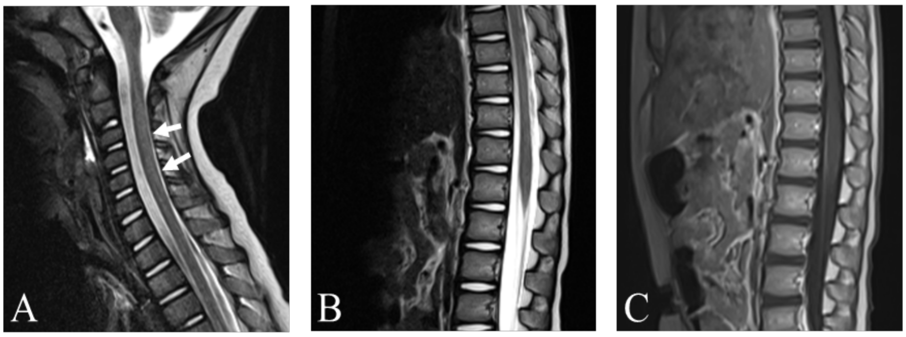


Select findings 8 months after presentation: Selected T2 axial (A, B), T2 FLAIR axial (C) and T2 FLAIR coronal (D) MR head images show ongoing deep white matter and periventricular high signal (arrows), particularly in the centrum semiovale and frontoparietal white matter, but also in the temporo-occipital lobes and cerebellum. Evidence of progressive cerebral atrophy with frontotemporal predominance (arrowheads).

White matter signal changes are non-specific – while they can be seen in progressive demyelinating disease or mitochondrial pathology, they can also be seen in response to cortical atrophy.

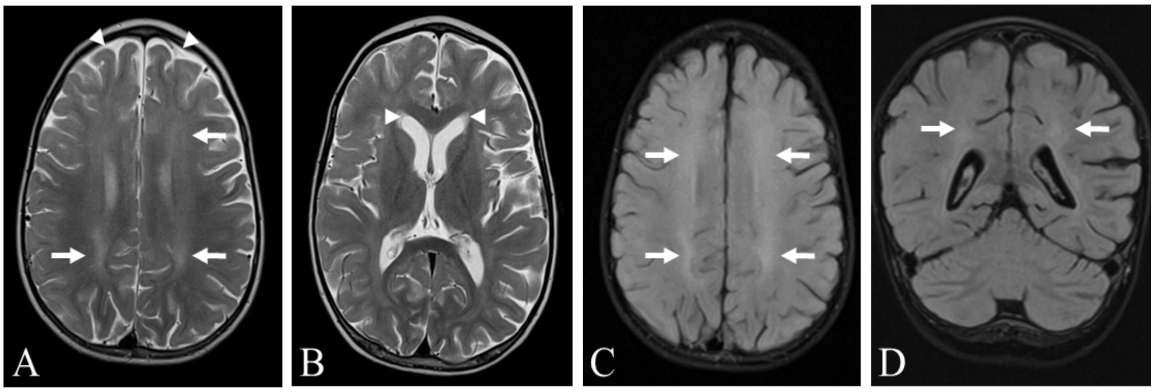

Patient 17

Initial MR at presentation was unremarkable. Later MR showed T2 hyperintensities in the supratentorial white matter.

Selected T2 axial images from a follow up MR (A, B) demonstrate frontotemporal predominant cerebral atrophy with associated prominence of the frontal extra-axial CSF spaces (arrows).

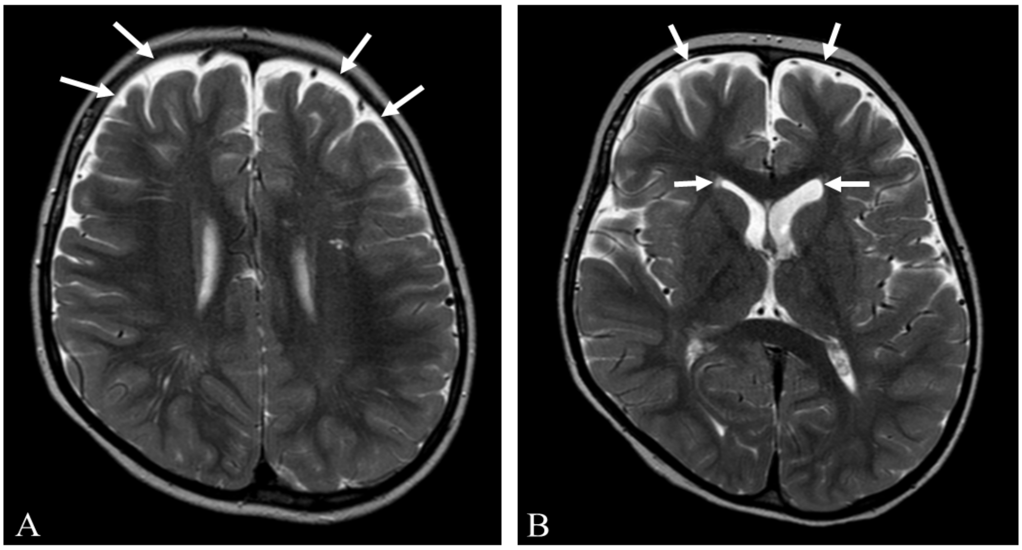

##### 3.2 Table S6. RCC1 variant details

| **Coding** | **Protein** | **Family ID** | **gnomAD v4** | **CADD v1.6** |
|---|---|---|---|---|
| c.127G>A | Gly43Ser | 1, 2 | Unseen<br>Gly43Alafs seen in 1x het(erozygote)<br>Gly43Gly seen in 8x het | 28.9 |
| C.209T>C | Val70Ala | 10 | Unseen | 27.2 |
| c.238G>A | Val80Met | 3 | Val80Met seen 47 het, no hom(ozygote) in 1,582,952 | 28.5 |
| c.280A>G | Asn94Asp | 4 | Unseen<br>Asn94Asp in 2x het | 28.6 |
| c.330G>C | Met110Ile | 5, 7, 8, 9, 10 | Met110Ile seen 28x het no hom in 1613954 | 11.55 |
| c.604G>A | Gly202Ser | 7 | Gly202Ser seen 2x het, no hom in 1,461,844<br>Gly202Arg seen 1x het | 28.8 |
| *C.767C>T* | *Ser256Phe* | *N/A* | *Unseen* | *23.5* |
| C.781G>A | Val261Met | 11, 12 | Val261Met seen in 6 het of 1,461,636 | 19.6 |
| c.1195C>T | Arg399Cys | 3, 6 | Arg399Cys seen 36x het, no hom in 1,613,730<br>Arg399His 11x het | 32 |

##### 3.3 Figure S2: Structural modelling of Rcc1 variants

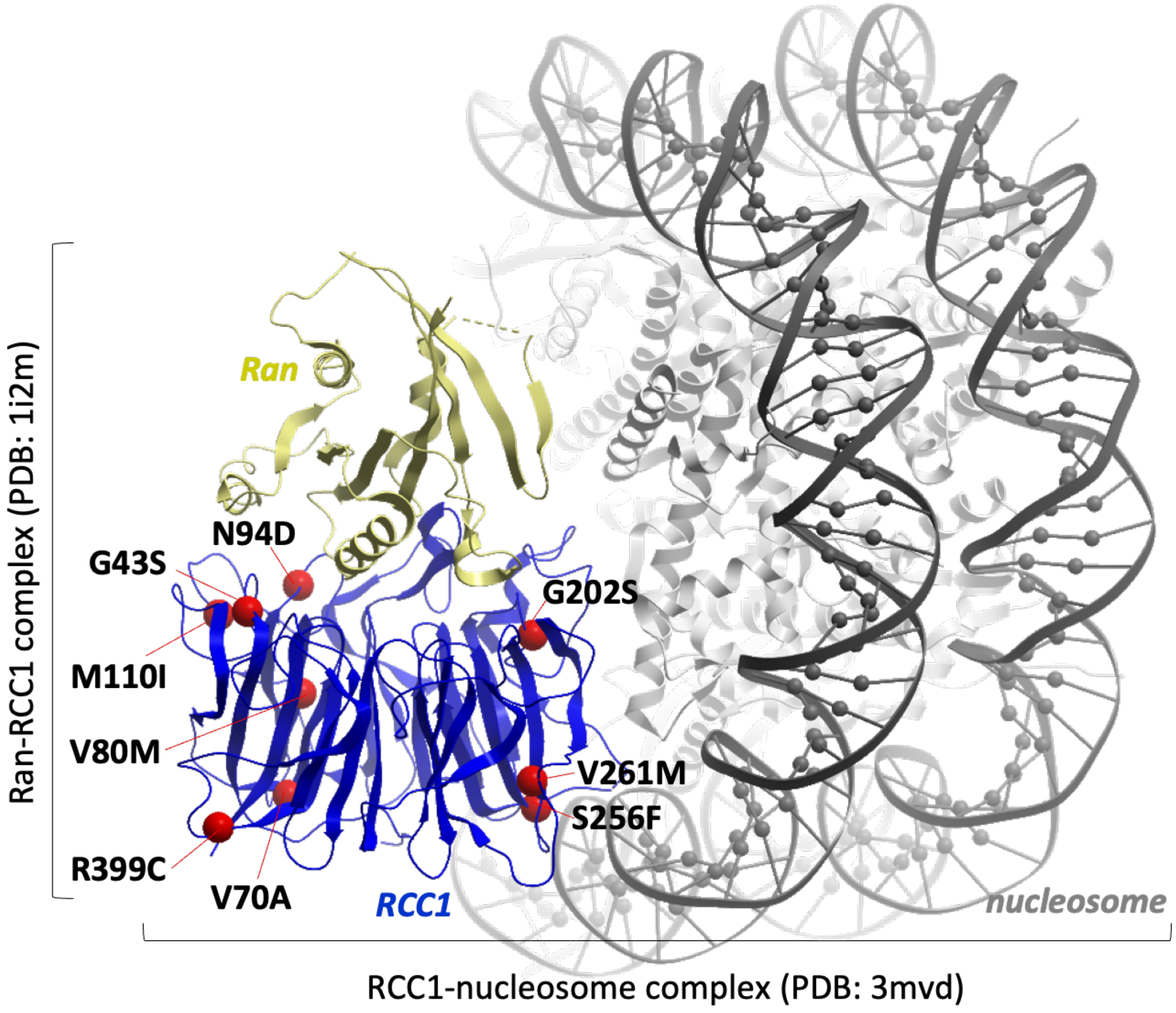


**Figure S2**. Modelling variant location using structure of Rcc1.

**3.4 Figure S3: Recombinant expression of Rcc1 mutant proteins.**

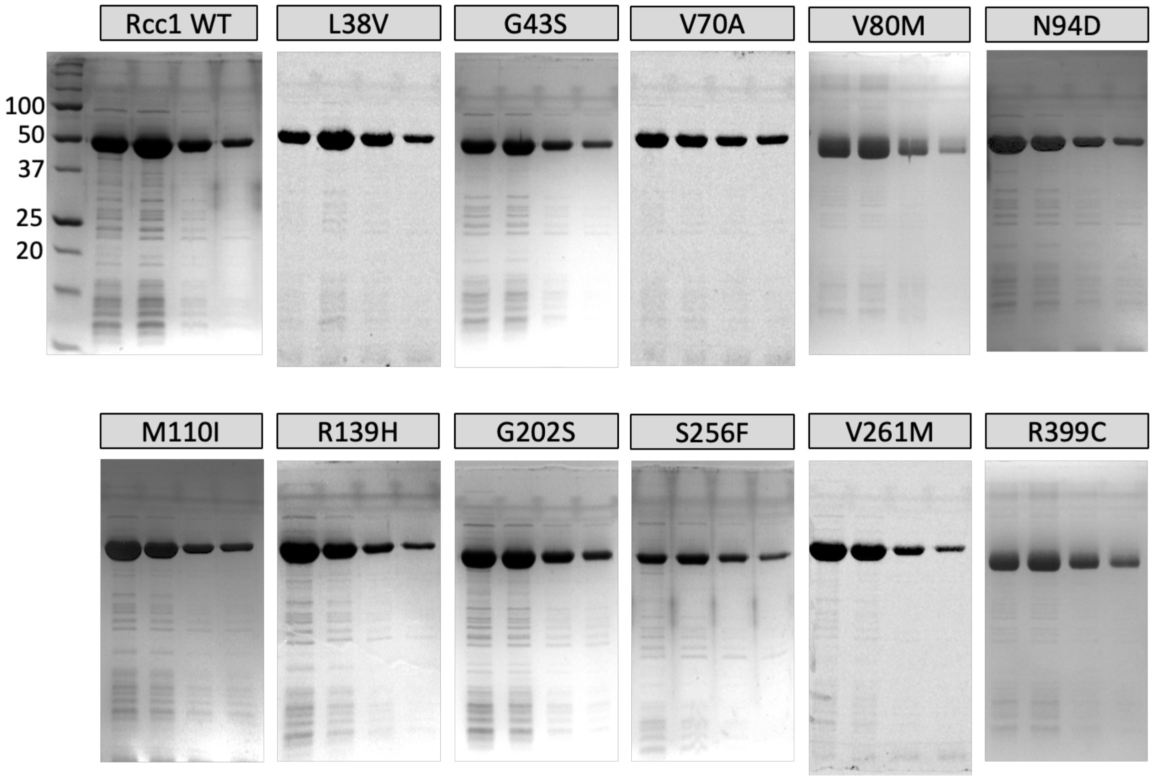


**Figure S3.** Purification of recombinant Rcc1 mutant proteins. Four 1 mL fractions were eluted, and 10µl eluate of each fraction was subjected to SDS-PAGE, followed by ReadyBlue staining.

#### 3.5 Figure S4: Thermal stability assay response curves

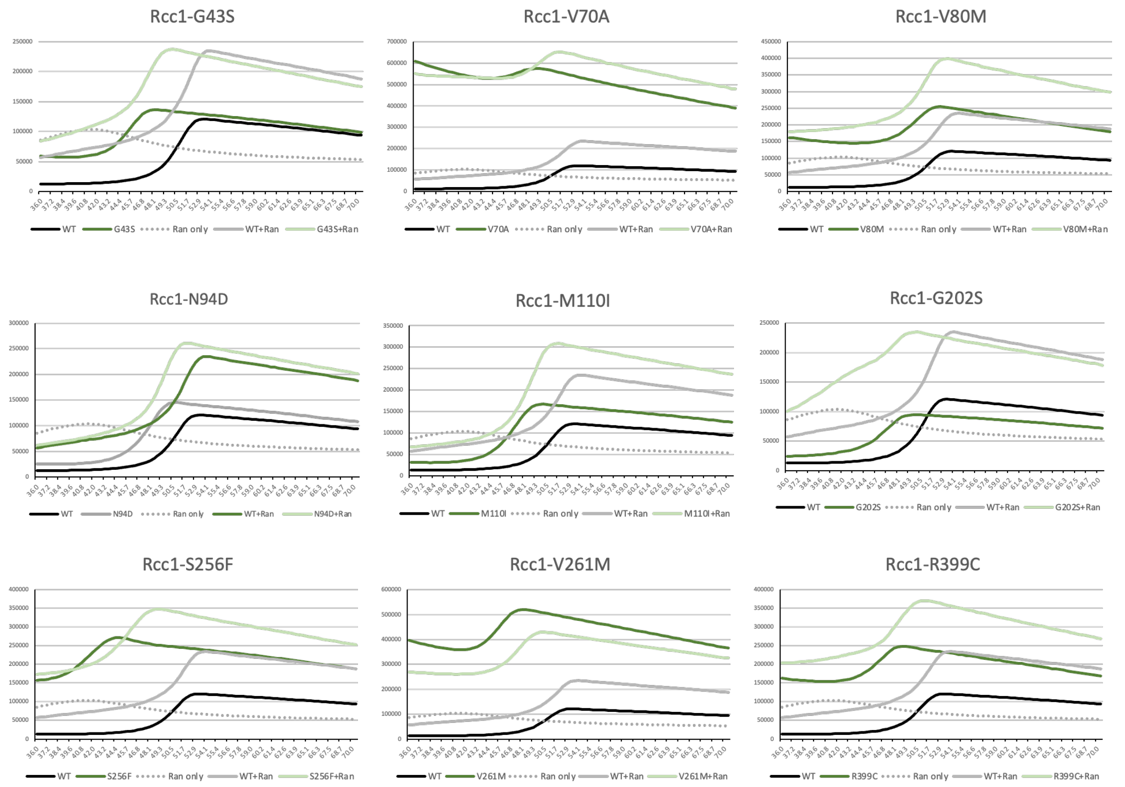


**Figure S4**. Protein thermal shift plots of Rcc1 mutants. Temperature plotted against fluorescence. $T_m$ determined by the temperature at which maximum fluorescence is reached.

##### 3.6 Figure S5: Western blotting of Rcc1 in patient fibroblast lysates

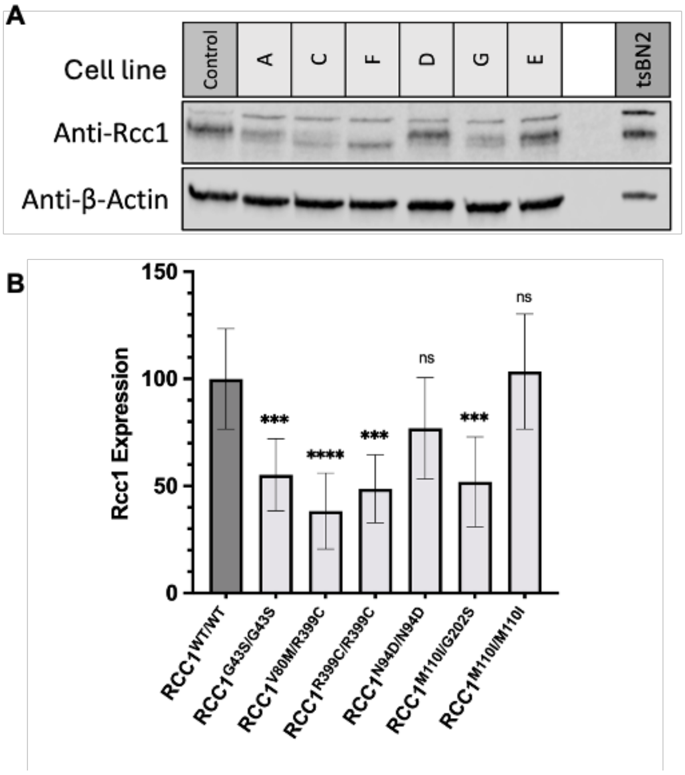


**Figure S5.** Western blotting of patient-derived fibroblast and tsBN2 whole-cell lysates. (A) Immunodetection of Rcc1 and β-Actin of control fibroblasts and six patient-derived fibroblasts, and the tsBN2 cell line. Cell lines: A, Patient 1; C, Patient 9; F, Patient 16; D, Patient 13; G, Patient 17; E, Patient 14. (B) Mean β-Actin-normalized Rcc1 fibroblast expression ± s.d. (n=3). Data were subjected to Dunnett's multiple comparisons test, *p<0.05, **p<0.01, ***p<0.001, ****p<0.0001, ns non-significant.


#### 5. Supplementary Acknowledgments and Institutional Support


A.B. is supported by a Wellcome PhD Training Fellowship for Clinicians and the 4Ward North PhD Programme for Health Professionals (223521/Z/21/Z).

P.L. is supported by Ministry of Health of the Czech Republic, grant. no: NW24-04-00349.

R.H. is supported by the Wellcome Discovery Award (226653/Z/22/Z), the Medical Research Council (UK) (MR/V009346/1), the Addenbrookes Charitable Trust (G100142), the Hereditary Neuropathy Foundation, the Stoneygate Trust, the Lily Foundation, Ataxia UK, Action for AT, the Muscular Dystrophy UK, the LifeArc Centre to Treat Mitochondrial Diseases (LAC-TreatMito) and the UKRI/Horizon Europe Guarantee MSCA Doctoral Network Programme (Project 101120256: MMM). This research was supported by the NIHR Cambridge Biomedical Research Centre (BRC-1215-20014). The views expressed are those of the authors and not necessarily those of the NIHR or the Department of Health and Social Care.

H.L. receives support from the Canadian Institutes of Health Research (CIHR) for Foundation Grant FDN-167281 (Precision Health for Neuromuscular Diseases), Transnational Team Grant ERT-174211 (ProDGNE) and Network Grant OR2-189333 (NMD4C), from the Canada Foundation for Innovation (CFI-JELF 38412), the Canada Research Chairs program (Canada Research Chair in Neuromuscular Genomics and Health, 950-232279), the European Commission (Grant # 101080249) and the Canada Research Coordinating Committee New Frontiers in Research Fund (NFRFG-2022-00033) for SIMPATHIC, and from the Government of Canada Canada First Research Excellence Fund (CFREF) for the Brain-Heart Interconnectome (CFREF-2022-00007).

RWT is funded by the Wellcome Centre for Mitochondrial Research (203105/Z/16/Z), the Mitochondrial Disease Patient Cohort (UK) (G0800674), the Medical Research Council (MR/W019027/1), the Lily Foundation, the Pathological Society, the UK NIHR Biomedical Research Centre for Ageing and Age-related disease award to the Newcastle upon Tyne Foundation Hospitals NHS Trust, LifeArc and the UK NHS Highly Specialised Service for Rare Mitochondrial Disorders of Adults and Children.

R.D.S.P. is funded by The Lily Foundation, Muscular Dystrophy UK (MDUK), and a seedcorn award from the Rosetrees Trust and Stoneygate Foundation. R.D.S.P. is supported by a Medical Research Council (UK) Transition Support award (MR/X02363X/1), Medical Research Council (UK) award MC_PC_21046 to establish a National Mouse Genetics Network Mitochondria Cluster (MitoCluster), and the LifeArc Centre to Treat Mitochondrial Diseases (LAC-TreatMito).
